# Causal relationship of polyunsaturated fatty acids with mental disorders: a systematic review and meta-analysis

**DOI:** 10.1101/2025.02.16.25322383

**Authors:** Yiying Yao, Xiaopeng Yang, Junwei Yan, Hongran Lv, Zihan Yue, Jiayao Yu, Mengfei Ye, Xiuqin Lin, Chao Qian, Huafang Zhang, Zheng Liu

**Author notes:** Correspondence: Mengfei Ye,; No.1234 Shengli West Road, Yuecheng District, Shaoxing 312000, Zhejiang, China Zheng Liu,; No.900 Chengnan Avenue, Yuecheng District, Shaoxing 312000, Zhejiang, China. These authors contributed equally to this work (Yiying Yao, Xiaopeng Yang, Junwei Yan).

## Abstract

Mental disorders (MDs) are emerging as a significant threat to global health, with their intricate pathogenesis making the discovery of effective treatments a challenging endeavor. Currently, nutritional intervention, especially polyunsaturated fatty acids (PUFAs), has garnered widespread attention. This review aims to explore the potential causal relationship between PUFAs and MDs using Mendelian randomization (MR) meta-analysis. The study employed two-sample MR technology to analyze the association between PUFAs (including omega-3, omega-3%, omega-6, omega-6%, and the omega-6:omega-3 ratio) and MDs. It utilized data from genome-wide association studies to assess the role of PUFAs in 12 major MDs. The MR analysis revealed a causal link between genetically predicted omega-3 and MDs such as obsessive-compulsive disorder, bipolar disorder, schizophrenia, and major depressive disorder. Omega-3% showed protective effects against certain diseases, including emotional personality disorder. Meanwhile, the inverse correlation between genetically predicted omega-6 and the risk of attention deficit/hyperactivity disorder, as well as the association between a high omega-6:omega-3 ratio and increased risk of depression and other mood disorders, were also supported by meta-analyses. These findings suggest that high levels of omega-3 and omega-3% may reduce the risk of MDs, while a high omega-6:omega-3 ratio may increase the risk. The study highlights the potential of PUFAs, particularly omega-3, in the prevention and treatment of MDs, while also noting the complex interactions between omega-3 and omega-6 and their impact on MDs, which necessitates further research. These findings provide a scientific basis for future clinical trials and the development of dietary intervention measures.

## 1 Introduction

Mental disorders (MDs) are a collection of syndromes marked by notable disruptions in cognitive function, emotional regulation, and behavior, indicative of underlying mental, biological, or developmental irregularities ^1^. Data from a 2023 survey revealed that the lifetime prevalence of MDs stands at 28.6% for male and 29.8% for female, with the initial onset typically occurring around the age of 15, a critical period that can significantly impact the early stages of an individual’s life cycle ^2^. The majority of MDs are preventable and treatable through professional psychological support, routine health screenings ^3^, and the administration of antipsychotic medications ^4^. Unfortunately, a concerning 75% of patients face debilitating side effects that not only degrade their quality of life but also contribute to social stigmatization and, in the most severe cases, lead to relapses or even fatal consequences. Moreover, the adherence rate to medication regimens is distressingly low, thereby undermining the effectiveness of long-term treatment strategies ^5^.

In light of these challenges, it is imperative to bolster preventive initiatives aimed at reducing the incidence of MDs. Empirical research consistently points to the benefits of healthy dietary practices and purposeful nutritional interventions in enhancing both the physical and psychological well-being of individuals ^6^. Furthermore, comprehensive meta-analyses have shown that these interventions could significantly reduce the risk of metabolic syndrome among those suffering from severe psychiatric conditions ^7^.

Polyunsaturated fatty acids (PUFAs), including omega-3 and omega-6 fatty acids, are pivotal dietary nutrients that are integral to neurodevelopment, brain function, behavior, and mental health ^8; 9^. Abundant in brain tissue, these fatty acids participate in a spectrum of brain biological processes, including metabolism, neurotransmission, synaptogenesis, and inflammation ^10^. A growing body of research implicates alterations in PUFA levels and function in a spectrum of neuropsychiatric disorders, such as Alzheimer’s disease (AD), attention deficit/hyperactivity disorder (ADHD), autism spectrum disorder (ASD), bipolar disorder (BD), schizophrenia (SCZ), and personality disorders (PsD) ^11^. For instance, omega-3 supplementation has been shown to ameliorate mood disorders and exhibits antidepressant effects in clinical settings ^12^. Serials of observational studies and meta-analyses have explored the relationship between PUFAs and psychiatric disorders. Some studies suggest that omega-3 supplementation enhances cognitive performance, while others propose that such benefits are confined to individuals with omega-3 deficiencies ^13–16^. A recent umbrella review of meta-analyses indicates that elevated PUFA levels may mitigate symptoms or lower the risk of various MDs, including depression and AD, although the evidence is predominantly weak, underscoring the need for comprehensive data synthesis to elucidate these relationships ^17^. Despite clinical and epidemiological studies suggesting a correlation between PUFAs and neuropsychiatric disorders, establishing a direct causal link remains elusive. This uncertainty is attributed to the paucity of large-scale randomized controlled trials and the influence of confounding variables ^9^.

Mendelian randomization (MR) is a robust statistical approach that leverages genetic variation as an instrumental variable to establish causality, effectively side-stepping the confounding factors and the issue of reverse causation that often complicate observational studies^12^. While MR has been utilized to probe the potential causal link between PUFAs and the risk of various MDs, the findings have been inconclusive. For instance, some studies propose a protective effect of omega-3s in major depressive disorder (MDD) ^18^, yet others have not found such associations to be statistically significant^19^. By integrating MR with meta-analyses, we can harness the extensive genetic variation data from genome-wide association studies (GWAS) to evaluate causal relationships on an expansive scale^20^.

In the present study, we aim to conduct a comprehensive MR analysis, treating PUFAs as the exposure and MDs as the outcomes. We will also undertake a meta-analysis of MR studies to determine the causal role of PUFAs in MDs and to assess the impact of varying types and ratios of PUFAs. Our research is designed to uncover the causal dynamics between PUFAs and MDs, with the ultimate goal of providing a solid theoretical framework for the clinical application of PUFAs in both the prevention and treatment of MDs.

## 2 Methods

This systematic review and meta-analysis was conducted in accordance with the preferred items in the PRISMA guidelines^21^. The corresponding programme for this systematic evaluation has been registered in the International Register of Prospective Systematic Evaluation (PROSPERO) and is publicly available online (CRD42024598472).

### 2.1 Study design

The two-sample MR study was designed to investigate a potential unidirectional causal pathway between PUFAs and MDs. In these analyses, PUFAs were considered as the exposure variable to determine if they exert a causal influence on MDs. We utilized summary-level data from large-scale, non-overlapping GWAS for all exposure and outcome variables. To evaluate the impact of different types of PUFAs, we examined five distinct PUFA exposures: omega-3, omega-3 percentage (omega-3%), omega-6, omega-6 percentage (omega-6%), and the ratio of omega-6 to omega-3 (omega-6:omega-3). In our study, we utilized an outcome range approach to evaluate the potential causal links between PUFAs and a spectrum of 12 MDs, along with their respective subgroups. Our analysis encompassed a variety of conditions, including depression, MDD, anxiety, post-traumatic stress disorder (PTSD), BD, SCZ, ADHD, mania, obsessive-compulsive disorder (OCD), ASD, PsD and AD.

### 2.2 GWAS data Sources

#### 2.2.1 Polyunsaturated Fatty Acids

Summary statistics for omega-3, omega-3%, omega-6, omega-6%, and omega-6:omega-3 are readily available on the IEU Open GWAS database. To ensure genetic similarity between the two samples, we have chosen the European population database as the focus of our study. The data IDs for all relevant records can be consulted in the Appendix.

#### 2.2.2 Data source for MDs

Diagnostic summary statistics for AD, ADHD, anxiety, ASD, BD, depression, mania, MDD, OCD, PTSD, SCZ and PsD were sourced from the IEU Open GWAS Project, UK Biobank, and FinnGen. Additional details regarding the data sources, sample sizes, and the strength of the genetic tools used in the GWAS dataset are provided in the appendix (Table S2). It is important to note that all original studies contributing to the GWAS dataset were conducted with ethical approval and with the informed consent of all participants.

### 2.3 Selection of Genetic Instruments

We developed a distinct set of genetic instruments for each exposure variable, focusing exclusively on single nucleotide polymorphisms (SNPs) that achieved genome-wide significance (p < 5×10^−8^). To ensure the independence of the genetic variants used as instruments, we employed a clumping procedure that excluded SNPs with a correlation coefficient (R^2^) greater than 0.001 and a linkage disequilibrium (LD) score greater than 10. To mitigate the influence of multicollinearity and over-correlated SNPs on our analysis, we conducted LD clumping with an aggregation distance parameter set at 10,000 kb. This methodological step ensured that the selected SNPs were independent, thereby enhancing the precision and reliability of our MR analysis. The complete list of SNPs utilized as instruments for each phenotype is detailed in Appendix 1 (Table S1).

### 2.4 Search strategy

Our literature search was conducted across electronic databases including PubMed, Embase, Web of science and the Cochrane Library. We focused on articles published prior to October 1, 2024, and restricted our search to studies involving human participants. There were no limitations imposed on the publication date. For further details on the search terms used, refer to Supplementary Table 1. In addition to electronic searches, we also performed manual searches and scrutinized the references of the included articles to identify additional relevant studies (backward manual search). From each study, we extracted the following information: author, publication year, country of origin, type of MR study, exposure data source(s), exposure variable(s), exposure sample size(s), number of single-nucleotide polymorphisms (SNPs) utilized, outcome data source(s), outcome variable(s), outcome sample size(s), MR analysis method, and results.

### 2.5 Inclusion and exclusion criteria

This systematic review and meta-analysis encompassed studies that: (1) investigated the associations between genetic liability to omega-3, omega-3%, omega-6, omega-6%, and omega-6:omega-3 with mental disorders, (2) employed MR methods involving human participants, and (3) were published in English. We excluded articles that were not MR studies, lacked a full-text, presented unrelated data, had unavailable data, or reported on the same dataset analyzed multiple times. For the purpose of quality assessment, the information extracted was guided by a template developed in accordance with the STROBE-MR guidelines ^22^, which aim to enhance the reporting of observational studies in epidemiology.

### 2.6 Statistical analysis

We evaluated the potential causal relationships between omega-3, omega-3%, omega-6, omega-6%, omega-6:omega-3 (as exposures) and MDs (as outcomes). In our primary analysis, we employed a random-effects instrumental variable weighted (IVW) regression model to synthesize effect estimates from multiple SNPs, using odds ratios (ORs) with their corresponding 95% confidence intervals (CIs).

For sensitivity analysis, we utilized a suite of robust MR methods to detect and rectify any potential breaches of MR assumptions. These methods included MR-Egger regression, weighted median, weighted mode, and MR-PRESSO. Furthermore, we conducted heterogeneity and multiplicity tests to validate the robustness of our findings. All MR statistical analyses were conducted using R (version 4.2.1) with the TwoSampleMR software package.

For the meta-analyses, we used STATA (version 12.0) and pooled the ORs from individual studies using a random-effects model. Forest plots were generated to graphically represent the pooled results ^23^. In our quantitative synthesis, we only included outcomes with at least two studies reporting non-overlapping data. In cases where two or more studies reported data from the same source, we included only the study with the largest participant count. Studies not included in the meta-analysis were still considered in the qualitative analysis. Heterogeneity was assessed using the I^2^ statistic, with thresholds set at 25–50% for mild, 50–75% for moderate, and >75% for severe heterogeneity ^24^. We visually inspected funnel plots to assess publication bias. Statistical significance was set at a two-sided P-value of less than 0.05.

## 3 Result

### 3.1 Literature Search Results

In this comprehensive study, we meticulously selected PUFAs, including omega-3, omega-3%, omega-6, omega-6%, and the omega-6:omega-3 ratio, from databases such as Finland, to serve as exposures. MDs were designated as the outcome measures. We conducted 419 de novo MR analyses to explore these relationships. Furthermore, we meticulously extracted 575 records from various databases. After a thorough deduplication process and a comprehensive review, 12 studies were identified as meeting our stringent eligibility criteria, including references^18; 19; 25–34^, which were subsequently reintegrated into our analysis. The meta-analysis encompassed an extensive dataset comprising 470 MR analyses (as depicted in **Figure 1**).

**Fig.1:**
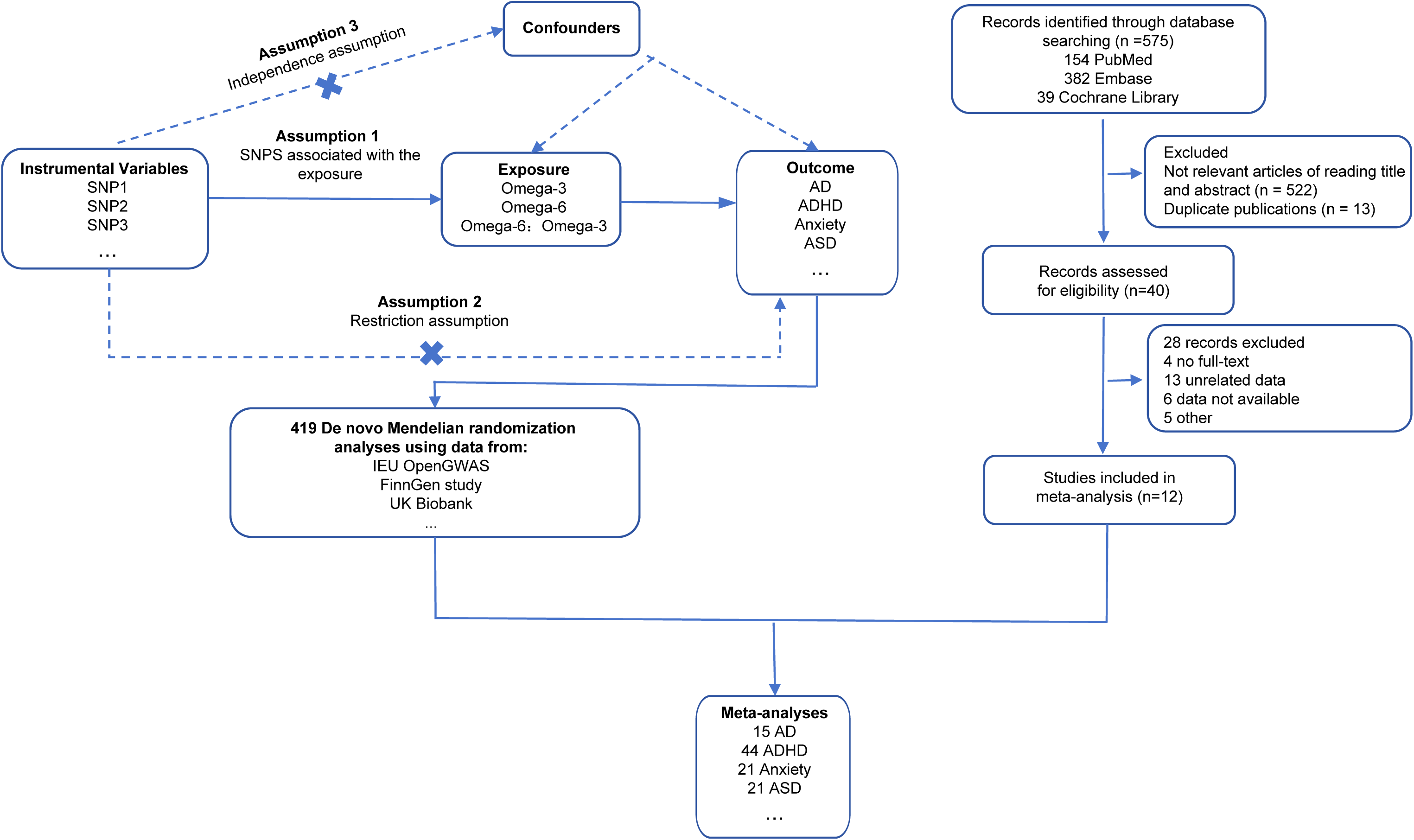
Flowchart of literature search, Mendelian randomization (MR), and meta-analysis. Abbreviations: AD, Alzheimer’s disease; ADHD, attention deficit/hyperactivity disorder; ASD, autism spectrum disorder; MR, Mendelian randomization.

The quality assessment and results of the included studies are meticulously detailed in **Table S4**. It is noteworthy that 42% of the studies acknowledged the presence of potential confounding factors, 75% reported relatively complete data information and 83% addressed sensitivity analysis. This study amalgamated large-scale datasets from a diverse array of countries, predominantly European, to investigate the potential causal links between the aforementioned exposure variables and MDs. Our exposure sample size is expansive, encompassing over 100,000 individuals from databases such as the IEU Open GWAS, UK Biobank, and the Canadian Longitudinal Study on Aging (CLSA) cohort. The SNPs featured in this paper underwent rigorous scrutiny to ensure their suitability as instrumental variables. We meticulously extracted results based on the inverse IVW method as the primary analysis objects for our meta-analysis. This rigorous and systematic approach ensures the robustness of our findings, contributing valuable insights into the complex relationship between PUFAs and MDs.

### 3.2 De novo MR analysis Results

The primary MR analysis results, utilizing the IVW method, are depicted in **Table S6**. Our findings indicate a protective effect of higher omega-3 levels against MDs. Specifically, there were seven MDD studies , two BD studies , and one study each for OCD and SCZ . When considering the percentage of omega-3 among PUFAs, data from two PsD-emotional studies and one anxiety study also suggested protection at higher omega-3 percentages.

However, results for MDD were inconsistent, showing five protective effects and four deleterious effects. Similarly, depression exhibited inconsistent outcomes. High levels of omega-6 were protective against ADHD, and high omega-6 percentages were beneficial for PsD. The distinction between the effects of high levels versus high percentages of omega-3 and omega-6 underscores the importance of examining the omega-6 to omega-3 ratio. Studies on MDD, depression, PsD-emotional, BD, and anxiety indicate that a high omega-6 to omega-3 ratio may be detrimental.

### 3.3 Meta-analysis Results

The results of our meta-analysis are presented in **Figures 2 and 3**. A comprehensive analysis (Figure 2A) indicated that higher levels of omega-3 (OR = 0.957, 95% CI: 0.947 to 0.968) and omega-3 as a percentage of total fatty acids (omega-3%; OR = 0.953, 95% CI: 0.942 to 0.964) were associated with a reduced incidence of MDs. Additionally, omega-6 as a percentage of total fatty acids (omega-6%; OR = 0.991, 95% CI: 0.983 to 1.000) demonstrated a weak protective effect against MDs. Conversely, a higher ratio of omega-6 to omega-3 (OR = 1.035, 95% CI: 1.025 to 1.045) correlated with an increased risk of MDs. However, the overall estimates for omega-6 showed no significant association with MDs. In Figure 2A and 2B, it was observed that overall, PUFAs had a positive effect on MDs.

**Fig.2:**
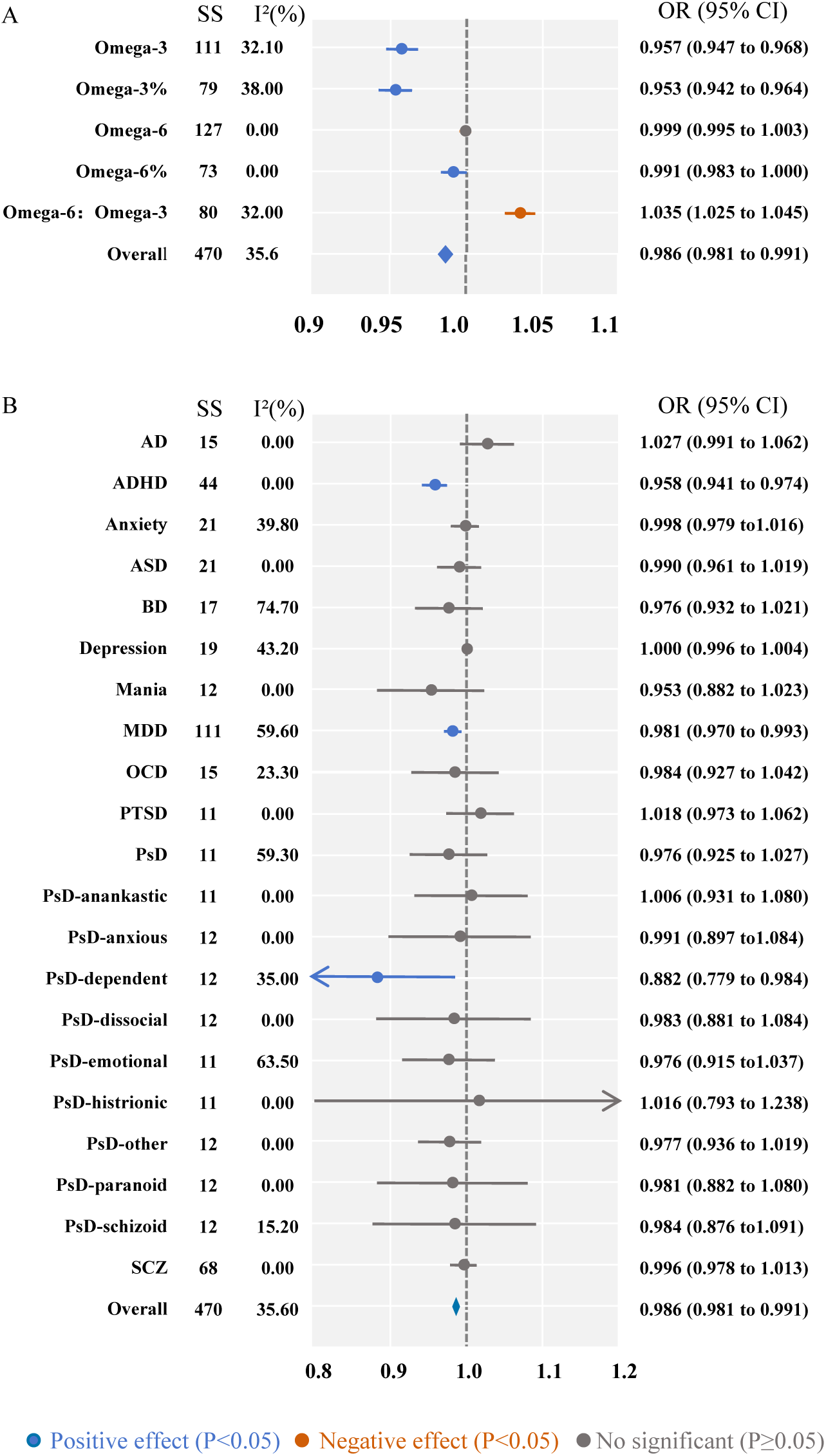
Forest plot of meta-analysis on MR results of polyunsaturated fatty acids (PUFAs) in mental disorders. (A) Meta-analysis of MR results for various PUFAs (omega-3, omega-3%, omega-6, omega-6%, omega-6:omega-3 ratio) in mental disorders. (B) Meta-analysis of MR results of PUFAs across different mental disorders. Abbreviations: AD, Alzheimer’s disease; ADHD, attention deficit hyperactivity disorder; ASD, autism spectrum disorder; BD, bipolar disorder; MDD, major depressive disorder; MR, Mendelian randomization; OR, odds ratio; OCD, obsessive-compulsive disorder; PTSD, post-traumatic stress disorder; PsD, personality disorder; PsD-anankastic, anankastic personality disorder; PsD-anxious, anxious personality disorder; PsD-dependent, dependent personality disorder; PsD-dissocial, dissocial personality disorder; PsD-emotional, emotionally unstable personality disorder; PsD-histrionic, histrionic personality disorder; PsD-other, mixed and other personality disorders; PsD-paranoid, paranoid personality disorder; PsD-schizoid, schizoid personality disorder; PUFAs, polyunsaturated fatty acids; SCZ, schizophrenia.

**Fig.3:**
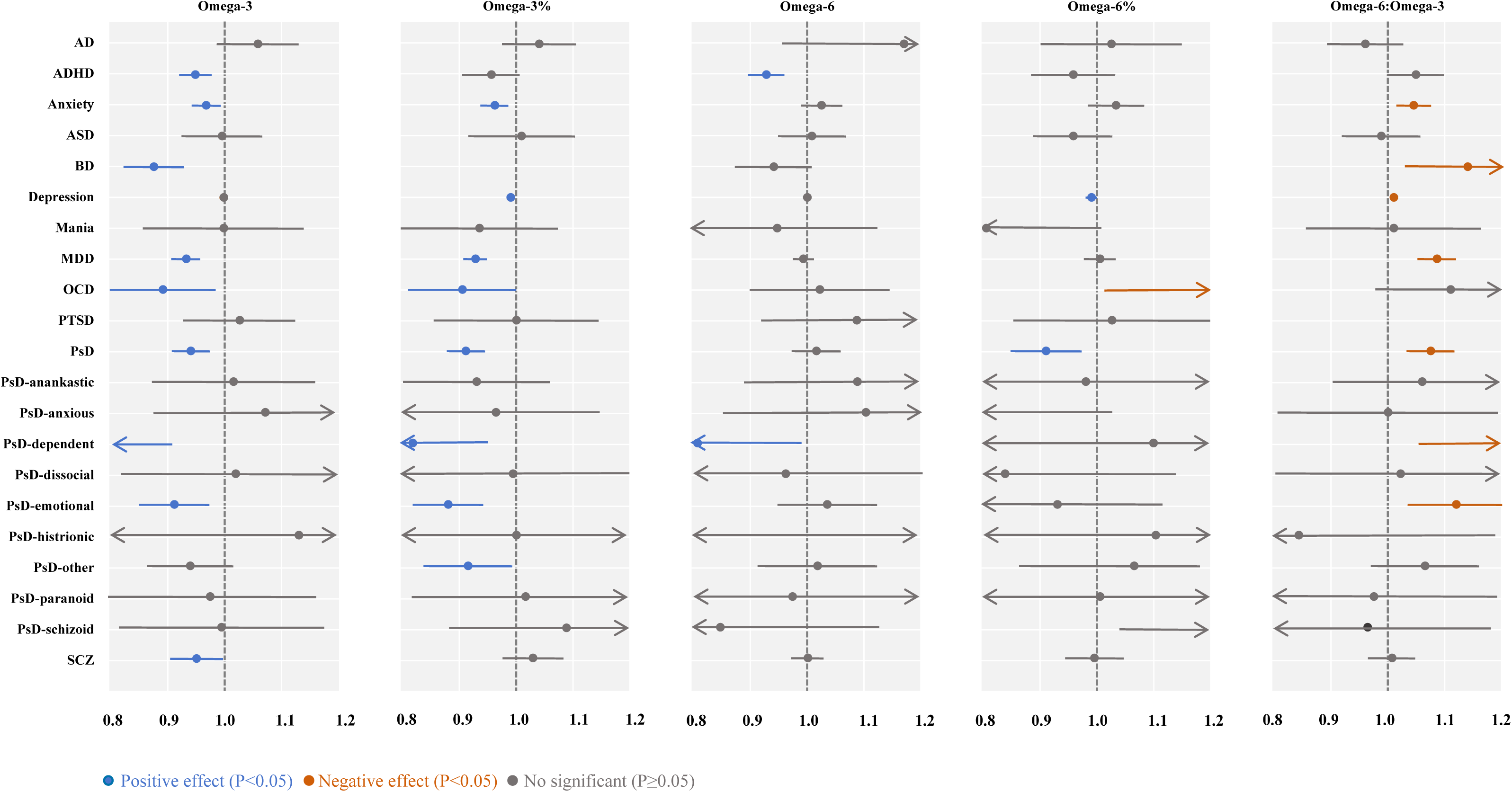
Forest plot of meta-analysis on subgroup analysis results. This plot reflects the subgroup results of a meta-analysis of MR estimates of causality for five PUFAs and various mental disorders. Abbreviations: AD, Alzheimer’s disease; ADHD, attention deficit hyperactivity disorder; ASD, autism spectrum disorder; BD, bipolar disorder; MDD, major depressive disorder; MR, Mendelian randomization; OR, odds ratio; OCD, obsessive-compulsive disorder; PTSD, post-traumatic stress disorder; PsD, personality disorder; PsD-anankastic, anankastic personality disorder; PsD-anxious, anxious personality disorder; PsD-dependent, dependent personality disorder; PsD-dissocial, dissocial personality disorder; PsD-emotional, emotionally unstable personality disorder; PsD-histrionic, histrionic personality disorder; PsD-other, mixed and other personality disorders; PsD-paranoid, paranoid personality disorder; PsD-schizoid, schizoid personality disorder; SCZ, schizophrenia.

**Fig.4:**
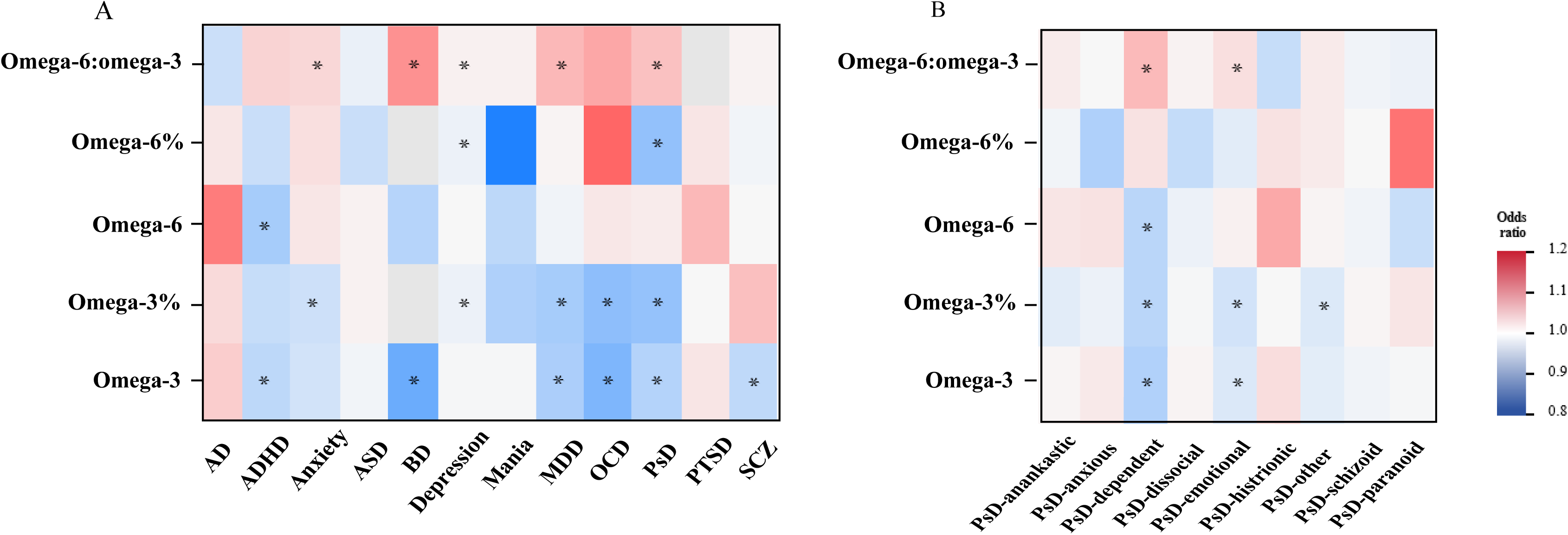
Heatmap of meta-analysis results for the effects of PUFAs on mental disorders. (A) Heatmap of meta-analysis results for the effects of PUFAs on 12 mental disorders. (B) Heatmap of meta-analysis results for the effects of PUFAs on personality disorder subgroups. Abbreviations: AD, Alzheimer’s disease; ADHD, attention deficit hyperactivity disorder; ASD, autism spectrum disorder; BD, bipolar disorder; MDD, major depressive disorder; OCD, obsessive-compulsive disorder; PTSD, post-traumatic stress disorder; PsD, personality disorder; PsD-anankastic, anankastic personality disorder; PsD-anxious, anxious personality disorder; PsD-dependent, dependent personality disorder; PsD-dissocial, dissocial personality disorder; PsD-emotional, emotionally unstable personality disorder; PsD-histrionic, histrionic personality disorder; PsD-other, mixed and other personality disorders; PsD-paranoid, paranoid personality disorder; PsD-schizoid, schizoid personality disorder; PUFAs, polyunsaturated fatty acids; SCZ, schizophrenia.

### 3.4 Subgroup analysis

In this study, significant heterogeneity was observed in the meta-analysis (Figure 2B). To explore the sources of this heterogeneity, a subgroup analysis was conducted to elucidate the relationship between PUFAs and MDs (**Figure 3** **and Table S5**).

Our analysis revealed that omega-3 fatty acids have significant protective effects against various MDs. Specifically, omega-3 significantly reduced the risk of ADHD, anxiety, BD, MDD, OCD, and SCZ. Additionally, high levels of omega-3 may be beneficial for PsD, particularly PsD-dependent and PsD-emotional subtypes.

Omega-3 as a percentage of total fatty acids (Omega-3%) acts as a protective factor against anxiety, depression , MDD, and OCD, and shows potential benefits for PsD and its subtypes: PsD-dependent, PsD-emotional, and PsD-other.

We found that omega-6 reduced the risk of ADHD and PsD-dependent. Omega-6 as a percentage of total fatty acids (Omega-6%) reduced the risk of depression and PsD.

Our results demonstrate that an increased omega-6 to omega-3 ratio elevates the risk of anxiety, BD, depression, and MDD, and is a risk factor for PsD , PsD-dependent, and PsD-emotional.

## 4. Discussion

### 4.1 Summary of evidence

In this comprehensive meta-analysis and systematic review, we analyzed 419 de novo MR studies and 12 published MR articles to explore the link between genetically predicted PUFAs and MDs. This study represents the first extensive evaluation of the causal effects of PUFAs on MDs using MR methodology. Our important findings include: 1) Genetic evidence suggests that higher omega-3 levels may reduce the risk of MDs, while a higher omega-6 to omega-3 ratio increases the risk of MDs. 2) High genetic omega-3 levels are associated with a reduced risk of ADHD, anxiety, BD, MDD, and PsD, both general and emotional subtypes. In contrast, a genetically higher omega-6 to omega-3 ratio is linked to an increased risk of these disorders.

Our study indicates that higher omega-3 levels protect against MDs, with docosahexaenoic acid (DHA) and eicosapentaenoic acid (EPA) being key components. DHA, abundant in the brain, aids in reducing brain inflammation and supports nerve development. EPA, though less abundant, influences neurotransmission and cell survival, thereby regulating mood and cognition^35^. However, omega-6, composed of linoleic acid and arachidonic acid, promotes inflammation contrary to omega-3’s anti-inflammatory effects. While no direct link between omega-6 levels and MDs was established, high omega-6 levels negatively impact MDs. The omega-6 to omega-3 ratio, not just levels, is a significant risk factor for MDs.

Our meta-analysis shows that higher omega-3 levels are linked to a reduced risk of various MDs, including ADHD, anxiety, BD, MDD, PsD, OCD, and SCZ. High omega-3 percentages also lower the risk of anxiety, depression, MDD, PsD, and OCD, suggesting omega-3 may benefit mental health. In ADHD, meta-analyses and studies affirm omega-3’s role in easing symptoms and lowering risk, presenting it as a viable non-pharmacological treatment option ^13; 17; 36^. Our MR studies reinforce this link, aligning with clinical findings. For anxiety, omega-3 appears beneficial, potentially by diminishing pro-inflammatory cytokines ^37^. Our meta-analysis indicates omega-3’s protective effects, recommending further exploration for anxiety treatment.

While a direct link between omega-3 and ASD isn’t established, disruptions in PUFA metabolism may play a role in ASD development ^38^. In BD and MDD, omega-3 deficiency could impair biosynthesis and cortical integrity, influencing disease progression ^39^. Our MR meta-analysis hints at a negative relationship between omega-3 and depression, though the efficacy of omega-3 supplementation for depression remains inconclusive ^40^. The impact of omega-3 on depression might be moderated by its ratio in supplements, depression severity, and individual characteristics ^41^. While beneficial for some, particularly with high EPA content, more research is needed to define omega-3’s role in treating MDs. Although not definitively linked to PsD, some evidence suggests omega-3’s positive influence on this disorder ^42^. Our MR findings further support a protective effect of omega-3 against OCD and SCZ, underscoring its potential in mental health management and indicating avenues for future research and clinical applications.

In the meta-analysis, we found that genetically associated omega-6 reduce the risk of ADHD and PsD-dependent, but are not statistically significantly associated with other MDs. Gamma-linolenic acid, an omega-6, shows potential as an adjunctive anti-inflammatory agent, which may improve ADHD symptoms ^43^. These findings highlight the multifaceted role of omega-6 in mental health, guiding future research towards elucidating the precise mechanisms through which omega-6 exerts its effects across various psychiatric disorders and synergistically impacts overall mental well-being.

The balance of omega-6 to omega-3 is crucial for mental health and implicated in chronic diseases. Modern Western diets often have an imbalanced ratio, with omega-6 exceeding omega-3, leading to health effects, especially in mental health disorders^44^. Imbalance in omega-3 to omega-6 ratios, particularly with high omega-6 intake like arachidonic acid, can displace essential EPA and DHA in cell membranes, leading to increased pro-inflammatory substances and disrupted cell functions. This imbalance is associated with a higher risk of MDs, potentially contributing to their development through effects on inflammation and neurotransmission ^45^. Recent insights into eicosanoid biochemistry have refined our view of omega-6, shifting away from labeling it solely as pro-inflammatory. New evidence suggests that higher omega-6 intake, including linoleic acid, doesn’t negatively impact inflammation or oxidative stress when omega-3 levels are adequate ^46^. The complex relationship between omega-3 intake, inflammation, and health emphasizes the need for further research into their effects on mental health.

### 4.2 Advantage and limitation

Our study’s strengths lie in its innovative methodology, robust data, and significant implications for mental health. By integrating MR with meta-analysis, we offer a fresh lens on the link between PUFAs and MDs. Leveraging extensive genetic data from GWAS, we bolster the reliability of our findings across 12 major MDs. Our results underscore the role of PUFAs, especially omega-3 and the omega-3 to omega-6 balance, in preventing and treating MDs, laying a foundation for future clinical trials and dietary interventions. With MDs’ high global prevalence, our findings could profoundly influence public health policies and dietary recommendations.

Certainly, the study has several limitations. Firstly, Two-sample MR is a powerful tool, but single-sample MR could confirm findings and assess confounding factors directly. Secondly, we could not evaluate the impact of specific fatty acid ratios (such as DHA and EPA) and combinations, which may be significant for clinical outcomes. Thirdly, most MR studies involve individuals of European descent, limiting the generalizability of PUFAs’ causal role in MDs to other populations. Therefore, more diverse and representative studies are needed to understand the global implications of PUFA intake and its relationship with mental health.

### 4.3 Conclusion

This MR study revealed a significant correlation: higher levels of omega-3 were associated with reduced risks of ADHD, BD, MDD, and various forms of PsD, whereas a higher omega-6 to omega-3 ratio was linked to an increased risk of these disorders. Additionally, genetically predicted higher percentages of omega-3 were found to be protective against depression, MDD, and multiple subtypes of PsD. The study provides limited evidence for the benefits of omega-6 and omega-6% in the context of mental disorders. These findings can guide decisions on the potential benefits and risks of PUFA intake. In conclusion, the association between PUFAs and mental health disorders underscores the need for further clinical trials to evaluate their therapeutic potential. Future interventions, including dietary adjustments and medications targeting polyunsaturated fatty acids, could pave the way for novel treatments aimed at preventing and managing mental health disorders.

## Acknowledgements

The authors would like to thank the FinnGen consortium, IEU Open GWAS database, UK Biobank for making summary-level data from this consortium publicly available.

## CRediT authorship contribution statement

Conceptualization, M.Y., Z.L. H.Z. and X. Y.; methodology, X.Y. and Y.Y; software, X.Y. and J. Y.; validation, J.Y.,Z.Y. and Y.Y.; formal analysis, X.Y. and H.L.; investigation, X.Y., Y.Y.; resources, X.Y., M.Y.; data curation, X.Y., Y.Y. J.Y.; writing—original draft preparation, X.Y.; writing—review and editing, M.Y., Z.L. H.Z. and C.Q.; visualization, X.Y., M.Y. and J.Y.; supervision, Z.L. and C.Q.; funding acquisition, M.Y. and X.L.. All authors have read and agreed to the published version of the manuscript.

## Data availability

No data were used for the research described in this article.

## Ethics approval and consent to participate

Not applicable.

## Consent for publication

All authors have agreed to publish this article.

## Abbreviations

AD: Alzheimer’s disease
ADHD: Attention deficit hyperactivity disorder
ASD: Autism spectrum disorder
BD: Bipolar disorder
CI: Confidence interval
DHA: Docosahexaenoic acid
EPA: Eicosapentaenoic acid
GWAS: Genome-wide association studies
IVW: Instrumental variable weighted
LD: Linkage disequilibrium
MDD: Major depressive disorder
MR: Mendelian randomization
MDs: Mental disorders
OCD: Obsessive-compulsive disorder
OR: Odds ratio
PsD: Personality disorder
PUFAs: Polyunsaturated fatty acids
PTSD: Post-traumatic stress disorder
PsD-anankastic: Anankastic personality disorder
PsD-anxious: Anxious personality disorder
PsD-dependent: Dependent personality disorder
PsD-dissocial: Dissocial personality disorder
PsD-emotional: Emotionally unstable personality disorder
PsD-histrionic: Histrionic personality disorder
PsD-other: Mixed and other personality disorders
PsD-paranoid: Paranoid personality disorder
PsD-schizoid: Schizoid personality disorder
SCZ: Schizophrenia
SNPs: Single nucleotide polymorphisms

