## supplemental 1 for "Causal relationship of polyunsaturated fatty acids with mental disorders: a systematic review and meta-analysis"

**Supplementary material**

[Table S1](#_Toc26885) [PRISMA 2020 Checklist	1](#_Toc26885)

[Table S2](#_Toc7868) [SNPs used as instrumental variables in the de novo Mendelian randomization analyses.	6](#_Toc7868)

[Table S3](#_Toc17839) [Overview of the used datasets.	29](#_Toc17839)

Table S1 PRISMA 2020 Checklist

| **Section and Topic** | **Item #** | **Checklist item** | **Location where item is reported** |
| --- | --- | --- | --- |
| **TITLE** | | |  |
| Title | 1 | Identify the report as a systematic review. | Title |
| **ABSTRACT** | | |  |
| Abstract | 2 | See the PRISMA 2020 for Abstracts checklist. | Abstract |
| **INTRODUCTION** | | |  |
| Rationale | 3 | Describe the rationale for the review in the context of existing knowledge. | Introduction, paragraphs 1 to 4 |
| Objectives | 4 | Provide an explicit statement of the objective(s) or question(s) the review addresses. | Introduction, paragraphs 5 |
| **METHODS** | | |  |
| Eligibility criteria | 5 | Specify the inclusion and exclusion criteria for the review and how studies were grouped for the syntheses. | Methods, paragraph 6 |
| Information sources | 6 | Specify all databases, registers, websites, organisations, reference lists and other sources searched or consulted to identify studies. Specify the date when each source was last searched or consulted. | Methods, paragraph 1 to 4,6 |
| Search strategy | 7 | Present the full search strategies for all databases, registers and websites, including any filters and limits used. | Methods, paragraph 6 |
| Selection process | 8 | Specify the methods used to decide whether a study met the inclusion criteria of the review, including how many reviewers screened each record and each report retrieved, whether they worked independently, and if applicable, details of automation tools used in the process. | Methods, paragraph 7 |
| Data collection process | 9 | Specify the methods used to collect data from reports, including how many reviewers collected data from each report, whether they worked independently, any processes for obtaining or confirming data from study investigators, and if applicable, details of automation tools used in the process. | Methods, paragraph 6 |
| Data items | 10a | List and define all outcomes for which data were sought. Specify whether all results that were compatible with each outcome domain in each study were sought (e.g. for all measures, time points, analyses), and if not, the methods used to decide which results to collect. | Methods, paragraph 6 |
|  | 10b | List and define all other variables for which data were sought (e.g. participant and intervention characteristics, funding sources). Describe any assumptions made about any missing or unclear information. | Methods, paragraph 6 |
| Study risk of bias assessment | 11 | Specify the methods used to assess risk of bias in the included studies, including details of the tool(s) used, how many reviewers assessed each study and whether they worked independently, and if applicable, details of automation tools used in the process. | Methods, paragraph 8 to 10 |
| Effect measures | 12 | Specify for each outcome the effect measure(s) (e.g. risk ratio, mean difference) used in the synthesis or presentation of results. | Methods, paragraph 8 to 10 |
| Synthesis methods | 13a | Describe the processes used to decide which studies were eligible for each synthesis (e.g. tabulating the study intervention characteristics and comparing against the planned groups for each synthesis (item #5)). | Methods, paragraph 8 to 10 |
|  | 13b | Describe any methods required to prepare the data for presentation or synthesis, such as handling of missing summary statistics, or data conversions. | Methods, paragraph 8 to 10 |
|  | 13c | Describe any methods used to tabulate or visually display results of individual studies and syntheses. | Methods, paragraph 8 to10 |
|  | 13d | Describe any methods used to synthesize results and provide a rationale for the choice(s). If meta-analysis was performed, describe the model(s), method(s) to identify the presence and extent of statistical heterogeneity, and software package(s) used. | Methods, paragraph 8 to 10 |
|  | 13e | Describe any methods used to explore possible causes of heterogeneity among study results (e.g. subgroup analysis, meta-regression). | Methods, paragraph 8 to 10 |
|  | 13f | Describe any sensitivity analyses conducted to assess robustness of the synthesized results. | Methods, paragraph 8 to 10 |
| Reporting bias assessment | 14 | Describe any methods used to assess risk of bias due to missing results in a synthesis (arising from reporting biases). | Methods, paragraph 8 to 10 |
| Certainty assessment | 15 | Describe any methods used to assess certainty (or confidence) in the body of evidence for an outcome. | Methods, paragraph 8 to 10 |
| **RESULTS** | | |  |
| Study selection | 16a | Describe the results of the search and selection process, from the number of records identified in the search to the number of studies included in the review, ideally using a flow diagram. | Results, paragraph 1 |
|  | 16b | Cite studies that might appear to meet the inclusion criteria, but which were excluded, and explain why they were excluded. | Results, paragraph 1 |
| Study characteristics | 17 | Cite each included study and present its characteristics. | Results, paragraph 2 |
| Risk of bias in studies | 18 | Present assessments of risk of bias for each included study. | Results, paragraph 2 |
| Results of individual studies | 19 | For all outcomes, present, for each study: (a) summary statistics for each group (where appropriate) and (b) an effect estimate and its precision (e.g. confidence/credible interval), ideally using structured tables or plots. | Results, paragraph 3 to 10 |
| Results of syntheses | 20a | For each synthesis, briefly summarise the characteristics and risk of bias among contributing studies. | Results, paragraph 3 to 10 |
|  | 20b | Present results of all statistical syntheses conducted. If meta-analysis was done, present for each the summary estimate and its precision (e.g. confidence/credible interval) and measures of statistical heterogeneity. If comparing groups, describe the direction of the effect. | Results, paragraph 3 to 10 |
|  | 20c | Present results of all investigations of possible causes of heterogeneity among study results. | Results, paragraph 3 to 10 |
|  | 20d | Present results of all sensitivity analyses conducted to assess the robustness of the synthesized results. | Results, paragraph 3 to 10 |
| Reporting biases | 21 | Present assessments of risk of bias due to missing results (arising from reporting biases) for each synthesis assessed. | Results, paragraph 3 to 10 |
| Certainty of evidence | 22 | Present assessments of certainty (or confidence) in the body of evidence for each outcome assessed. | Results, paragraph 3 to 10 |
| **DISCUSSION** | | |  |
| Discussion | 23a | Provide a general interpretation of the results in the context of other evidence. | Discussions, paragraphs 1 to 6 |
|  | 23b | Discuss any limitations of the evidence included in the review. | Discussions, paragraphs 8 |
|  | 23c | Discuss any limitations of the review processes used. | Discussions, paragraphs 8 |
|  | 23d | Discuss implications of the results for practice, policy, and future research. | Discussions, paragraphs 7 |
| **OTHER INFORMATION** | | |  |
| Registration and protocol | 24a | Provide registration information for the review, including register name and registration number, or state that the review was not registered. | Methods, paragraph 1 |
|  | 24b | Indicate where the review protocol can be accessed, or state that a protocol was not prepared. | Methods, paragraph 1 |
|  | 24c | Describe and explain any amendments to information provided at registration or in the protocol. | Methods, paragraph 1 |
| Support | 25 | Describe sources of financial or non-financial support for the review, and the role of the funders or sponsors in the review. | Funding |
| Competing interests | 26 | Declare any competing interests of review authors. | Declaration of Competing Interest |
| Availability of data, code and other materials | 27 | Report which of the following are publicly available and where they can be found: template data collection forms; data extracted from included studies; data used for all analyses; analytic code; any other materials used in the review. | Supplementary information is available at MF's website |

Table S2 SNPs used as instrumental variables in the de novo Mendelian randomization analyses.

| **Exposure** | **ID** | **SNP** | **Position** | **A1** | **A2** | **Beta** | **Eaf** | **Chr** | **SE** | **P** | **Samplesize** | **F** |
| --- | --- | --- | --- | --- | --- | --- | --- | --- | --- | --- | --- | --- |
| Omega-3 | ebi-a-GCST90092931 | rs660240 | 109817838 | C | T | 0.0370395 | 0.784707 | 1 | 0.00481655 | 1.50E-14 | 115006 | 59.1368 |
|  | ebi-a-GCST90092931 | rs12037485 | 2332391 | T | C | 0.0222781 | 0.460471 | 1 | 0.00396786 | 2.00E-08 | 115006 | 31.5242 |
|  | ebi-a-GCST90092931 | rs583609 | 62916796 | C | T | -0.0711993 | 0.352508 | 1 | 0.00414084 | 2.90E-66 | 115006 | 295.6477 |
|  | ebi-a-GCST90092931 | rs6547409 | 21190209 | T | C | -0.0565011 | 0.051259 | 2 | 0.0090697 | 4.70E-10 | 115006 | 38.8086 |
|  | ebi-a-GCST90092931 | rs11681659 | 136820960 | T | C | -0.0264162 | 0.716465 | 2 | 0.00436662 | 1.50E-09 | 115006 | 36.5974 |
|  | ebi-a-GCST90092931 | rs312939 | 21409548 | A | G | 0.034251 | 0.752467 | 2 | 0.00458283 | 7.80E-14 | 115006 | 55.8572 |
|  | ebi-a-GCST90092931 | rs1260326 | 27730940 | C | T | -0.0830756 | 0.604008 | 2 | 0.00404264 | 7.70E-94 | 115006 | 422.2959 |
|  | ebi-a-GCST90092931 | rs4860987 | 69491284 | T | A | 0.0450329 | 0.258612 | 4 | 0.00478075 | 4.50E-21 | 115006 | 88.7294 |
|  | ebi-a-GCST90092931 | rs11242109 | 131677047 | T | G | 0.0234291 | 0.479035 | 5 | 0.00395796 | 3.20E-09 | 115006 | 35.0403 |
|  | ebi-a-GCST90092931 | rs6882345 | 156397673 | A | G | 0.0274277 | 0.63285 | 5 | 0.00409997 | 2.20E-11 | 115006 | 44.7525 |
|  | ebi-a-GCST90092931 | rs2247056 | 31265490 | C | T | 0.0356403 | 0.703307 | 6 | 0.00432842 | 1.80E-16 | 115006 | 67.7991 |
|  | ebi-a-GCST90092931 | rs10455872 | 161010118 | G | A | -0.0617415 | 0.078987 | 6 | 0.00733401 | 3.80E-17 | 115006 | 70.8715 |
|  | ebi-a-GCST90092931 | rs117733303 | 160922870 | G | A | -0.110595 | 0.018512 | 6 | 0.0146728 | 4.80E-14 | 115006 | 56.8126 |
|  | ebi-a-GCST90092931 | rs9273217 | 32613747 | G | T | -0.0245669 | 0.318014 | 6 | 0.00448023 | 4.20E-08 | 115006 | 30.0677 |
|  | ebi-a-GCST90092931 | rs4000713 | 25990597 | A | G | -0.028225 | 0.295416 | 7 | 0.0043421 | 8.00E-11 | 115006 | 42.2540 |
|  | ebi-a-GCST90092931 | rs62466318 | 73042085 | T | C | -0.0716979 | 0.204201 | 7 | 0.00492892 | 6.20E-48 | 115006 | 211.5969 |
|  | ebi-a-GCST90092931 | rs112875651 | 126506694 | A | G | -0.0874051 | 0.392337 | 8 | 0.00410836 | 1.90E-100 | 115006 | 452.6229 |
|  | ebi-a-GCST90092931 | rs9987289 | 9183358 | G | A | 0.0544031 | 0.909161 | 8 | 0.00688493 | 2.70E-15 | 115006 | 62.4379 |
|  | ebi-a-GCST90092931 | rs10096633 | 19830921 | T | C | -0.0418412 | 0.123902 | 8 | 0.00599603 | 3.00E-12 | 115006 | 48.6946 |
|  | ebi-a-GCST90092931 | rs1800978 | 107665978 | G | C | -0.0387593 | 0.123988 | 9 | 0.00603019 | 1.30E-10 | 115006 | 41.3133 |
|  | ebi-a-GCST90092931 | rs3039101 | 5246177 | TGTA | T | -0.0352443 | 0.153601 | 10 | 0.00550396 | 1.50E-10 | 115006 | 41.0041 |
|  | ebi-a-GCST90092931 | rs7924036 | 65191645 | T | G | 0.0222276 | 0.504218 | 10 | 0.0039569 | 1.90E-08 | 115006 | 31.5555 |
|  | ebi-a-GCST90092931 | rs55891451 | 96728169 | C | A | 0.033166 | 0.20172 | 10 | 0.00494622 | 2.00E-11 | 115006 | 44.9613 |
|  | ebi-a-GCST90092931 | rs145786300 | 61406089 | A | G | -0.150855 | 0.011993 | 11 | 0.0187118 | 7.50E-16 | 115006 | 64.9962 |
|  | ebi-a-GCST90092931 | rs143355652 | 61453822 | T | C | -0.153201 | 0.010466 | 11 | 0.0198593 | 1.20E-14 | 115006 | 59.5107 |
|  | ebi-a-GCST90092931 | rs12226389 | 61823630 | C | T | -0.0535751 | 0.185805 | 11 | 0.00510749 | 9.70E-26 | 115006 | 110.0300 |
|  | ebi-a-GCST90092931 | rs2232143 | 60899701 | C | T | 0.104052 | 0.021879 | 11 | 0.0141105 | 1.70E-13 | 115006 | 54.3771 |
|  | ebi-a-GCST90092931 | rs174564 | 61588305 | G | A | -0.336931 | 0.347009 | 11 | 0.00412782 | 1.00E-200 | 115006 | 6662.5494 |
|  | ebi-a-GCST90092931 | rs964184 | 116648917 | C | G | -0.117565 | 0.867233 | 11 | 0.005805 | 3.40E-91 | 115006 | 410.1584 |
|  | ebi-a-GCST90092931 | rs145659493 | 61850279 | A | C | 0.107728 | 0.015853 | 11 | 0.0158266 | 1.00E-11 | 115006 | 46.3321 |
|  | ebi-a-GCST90092931 | rs149820547 | 61983775 | G | T | -0.0678421 | 0.042105 | 11 | 0.00988147 | 6.60E-12 | 115006 | 47.1363 |
|  | ebi-a-GCST90092931 | rs141469619 | 116714293 | G | A | 0.117162 | 0.010139 | 11 | 0.0208113 | 1.80E-08 | 115006 | 31.6939 |
|  | ebi-a-GCST90092931 | rs78689694 | 126234820 | C | G | 0.0326453 | 0.133362 | 11 | 0.00581875 | 2.00E-08 | 115006 | 31.4762 |
|  | ebi-a-GCST90092931 | rs11230829 | 61701898 | G | A | -0.103943 | 0.027785 | 11 | 0.0144233 | 5.70E-13 | 115006 | 51.9351 |
|  | ebi-a-GCST90092931 | rs673335 | 75450576 | C | T | -0.0684035 | 0.159752 | 11 | 0.00539262 | 7.20E-37 | 115006 | 160.9004 |
|  | ebi-a-GCST90092931 | rs7970695 | 121423376 | A | G | -0.0235143 | 0.620564 | 12 | 0.0040847 | 8.60E-09 | 115006 | 33.1393 |
|  | ebi-a-GCST90092931 | rs6602911 | 114547372 | T | C | 0.0231802 | 0.360074 | 13 | 0.00412043 | 1.80E-08 | 115006 | 31.6482 |
|  | ebi-a-GCST90092931 | rs139974673 | 44027885 | C | T | 0.118563 | 0.025916 | 15 | 0.0124678 | 1.90E-21 | 115006 | 90.4313 |
|  | ebi-a-GCST90092931 | rs261290 | 58678720 | C | T | -0.112905 | 0.654648 | 15 | 0.00416818 | 1.40E-161 | 115006 | 733.7252 |
|  | ebi-a-GCST90092931 | rs633695 | 58725839 | G | A | 0.0848151 | 0.292334 | 15 | 0.00436378 | 3.80E-84 | 115006 | 377.7641 |
|  | ebi-a-GCST90092931 | rs34663616 | 58569330 | A | C | 0.035253 | 0.137651 | 15 | 0.00586293 | 1.80E-09 | 115006 | 36.1545 |
|  | ebi-a-GCST90092931 | rs72789541 | 15127534 | A | T | -0.0834033 | 0.295973 | 16 | 0.00434225 | 3.20E-82 | 115006 | 368.9239 |
|  | ebi-a-GCST90092931 | rs35390787 | 15678414 | CA | C | 0.0223258 | 0.438254 | 16 | 0.00407642 | 4.30E-08 | 115006 | 29.9955 |
|  | ebi-a-GCST90092931 | rs16940904 | 44186063 | T | C | -0.0345078 | 0.226566 | 17 | 0.00474779 | 3.60E-13 | 115006 | 52.8265 |
|  | ebi-a-GCST90092931 | rs77960347 | 47109955 | G | A | 0.157178 | 0.013234 | 18 | 0.0172904 | 9.90E-20 | 115006 | 82.6368 |
|  | ebi-a-GCST90092931 | rs2187375 | 47172283 | G | A | 0.0524746 | 0.822879 | 18 | 0.00518371 | 4.40E-24 | 115006 | 102.4748 |
|  | ebi-a-GCST90092931 | rs12957708 | 47284353 | G | A | 0.0311923 | 0.14228 | 18 | 0.00568503 | 4.10E-08 | 115006 | 30.1044 |
|  | ebi-a-GCST90092931 | rs737338 | 11347657 | T | C | -0.0674466 | 0.035184 | 19 | 0.0107428 | 3.40E-10 | 115006 | 39.4171 |
|  | ebi-a-GCST90092931 | rs58542926 | 19379549 | T | C | -0.172929 | 0.074379 | 19 | 0.00754654 | 3.30E-116 | 115006 | 525.0974 |
|  | ebi-a-GCST90092931 | rs157592 | 45424514 | C | A | 0.0325574 | 0.185273 | 19 | 0.00522115 | 4.50E-10 | 115006 | 38.8837 |
|  | ebi-a-GCST90092931 | rs5112 | 45430280 | G | C | 0.0495704 | 0.533721 | 19 | 0.00425566 | 2.30E-31 | 115006 | 135.6785 |
|  | ebi-a-GCST90092931 | rs2288912 | 45449199 | G | C | 0.026507 | 0.496385 | 19 | 0.00396428 | 2.30E-11 | 115006 | 44.7087 |
|  | ebi-a-GCST90092931 | rs182611493 | 19458388 | G | A | -0.220029 | 0.012519 | 19 | 0.0190586 | 7.80E-31 | 115006 | 133.2840 |
|  | ebi-a-GCST90092931 | rs6129624 | 39167592 | A | G | -0.0242913 | 0.335232 | 20 | 0.0042636 | 1.20E-08 | 115006 | 32.4600 |
|  | ebi-a-GCST90092931 | rs117143374 | 40555561 | C | T | -0.03454 | 0.14225 | 21 | 0.00569242 | 1.30E-09 | 115006 | 36.8172 |
|  | met-d-Omega_3 | rs6693447 | 2330190 | G | T | 0.0229488 | 0.461686 | 1 | 0.00407772 | 4.80E-09 | 114999 | NA |
|  | met-d-Omega_3 | rs1167998 | 62931632 | A | C | 0.0713574 | 0.644674 | 1 | 0.00425038 | 3.60E-66 | 114999 | NA |
|  | met-d-Omega_3 | rs629301 | 109818306 | T | G | 0.0382887 | 0.778033 | 1 | 0.00488385 | 1.30E-14 | 114999 | NA |
|  | met-d-Omega_3 | rs1260326 | 27730940 | C | T | -0.0820592 | 0.60401 | 2 | 0.0041534 | 8.40E-88 | 114999 | NA |
|  | met-d-Omega_3 | rs11681659 | 136820960 | T | C | -0.0251255 | 0.716465 | 2 | 0.004486 | 2.00E-08 | 114999 | NA |
|  | met-d-Omega_3 | rs35135293 | 20363666 | T | C | -0.0208868 | 0.51675 | 2 | 0.00408348 | 3.90E-08 | 114999 | NA |
|  | met-d-Omega_3 | rs13424225 | 241214158 | T | G | 0.0221268 | 0.449809 | 2 | 0.00408591 | 2.20E-08 | 114999 | NA |
|  | met-d-Omega_3 | rs10184054 | 21203877 | G | C | -0.0361462 | 0.224107 | 2 | 0.00486548 | 5.60E-15 | 114999 | NA |
|  | met-d-Omega_3 | rs11563251 | 234679384 | T | C | 0.0349727 | 0.110601 | 2 | 0.00647476 | 3.20E-08 | 114999 | NA |
|  | met-d-Omega_3 | rs4860987 | 69491284 | T | A | 0.0463493 | 0.258609 | 4 | 0.00491057 | 1.20E-21 | 114999 | NA |
|  | met-d-Omega_3 | rs11242109 | 131677047 | T | G | 0.0240622 | 0.479016 | 5 | 0.0040658 | 2.40E-09 | 114999 | NA |
|  | met-d-Omega_3 | rs6882345 | 156397673 | A | G | 0.0288844 | 0.632863 | 5 | 0.00421159 | 1.90E-13 | 114999 | NA |
|  | met-d-Omega_3 | rs2394976 | 31311912 | T | G | -0.0461429 | 0.161648 | 6 | 0.00550837 | 1.20E-15 | 114999 | NA |
|  | met-d-Omega_3 | rs10455872 | 161010118 | G | A | -0.0629576 | 0.078988 | 6 | 0.00753435 | 2.80E-17 | 114999 | NA |
|  | met-d-Omega_3 | rs3129962 | 32379383 | A | G | -0.0392017 | 0.129765 | 6 | 0.00603734 | 1.80E-09 | 114999 | NA |
|  | met-d-Omega_3 | rs117733303 | 160922870 | G | A | -0.115945 | 0.018513 | 6 | 0.0150731 | 1.40E-15 | 114999 | NA |
|  | met-d-Omega_3 | rs62466318 | 73042085 | T | C | -0.0721329 | 0.204178 | 7 | 0.00506371 | 1.20E-45 | 114999 | NA |
|  | met-d-Omega_3 | rs73109460 | 44785800 | A | G | -0.0349475 | 0.123622 | 7 | 0.006216 | 9.20E-10 | 114999 | NA |
|  | met-d-Omega_3 | rs4000713 | 25990597 | A | G | -0.0288196 | 0.295408 | 7 | 0.00446039 | 1.00E-11 | 114999 | NA |
|  | met-d-Omega_3 | rs112875651 | 126506694 | A | G | -0.0873719 | 0.392346 | 8 | 0.00422001 | 3.50E-98 | 114999 | NA |
|  | met-d-Omega_3 | rs9987289 | 9183358 | G | A | 0.0566995 | 0.909151 | 8 | 0.00707191 | 3.20E-16 | 114999 | NA |
|  | met-d-Omega_3 | rs7819706 | 19844415 | G | A | -0.0396447 | 0.118291 | 8 | 0.00628878 | 1.80E-10 | 114999 | NA |
|  | met-d-Omega_3 | rs1800978 | 107665978 | G | C | -0.0373414 | 0.123991 | 9 | 0.00619432 | 5.20E-09 | 114999 | NA |
|  | met-d-Omega_3 | rs55891451 | 96728169 | C | A | 0.0341853 | 0.201728 | 10 | 0.00508075 | 4.60E-12 | 114999 | NA |
|  | met-d-Omega_3 | rs7924036 | 65191645 | T | G | 0.0233527 | 0.504205 | 10 | 0.00406452 | 5.50E-10 | 114999 | NA |
|  | met-d-Omega_3 | rs6601924 | 5247302 | C | T | 0.0350603 | 0.845765 | 10 | 0.00563873 | 8.50E-10 | 114999 | NA |
|  | met-d-Omega_3 | rs673335 | 75450576 | C | T | -0.0669996 | 0.159762 | 11 | 0.00554146 | 1.10E-34 | 114999 | NA |
|  | met-d-Omega_3 | rs12226389 | 61823630 | C | T | -0.050608 | 0.18582 | 11 | 0.00524839 | 1.10E-22 | 114999 | NA |
|  | met-d-Omega_3 | rs3018731 | 61248776 | G | A | -0.0353357 | 0.717549 | 11 | 0.00456603 | 2.00E-14 | 114999 | NA |
|  | met-d-Omega_3 | rs144018203 | 116916060 | C | G | 0.107134 | 0.010653 | 11 | 0.02049 | 4.20E-08 | 114999 | NA |
|  | met-d-Omega_3 | rs143355652 | 61453822 | T | C | -0.154138 | 0.010467 | 11 | 0.0204073 | 9.40E-14 | 114999 | NA |
|  | met-d-Omega_3 | rs174564 | 61588305 | G | A | -0.337094 | 0.347013 | 11 | 0.00424185 | 1.00E-200 | 114999 | NA |
|  | met-d-Omega_3 | rs964184 | 116648917 | C | G | -0.116637 | 0.867229 | 11 | 0.00596503 | 8.90E-87 | 114999 | NA |
|  | met-d-Omega_3 | rs11230829 | 61701898 | G | A | -0.102959 | 0.027786 | 11 | 0.0148212 | 3.40E-12 | 114999 | NA |
|  | met-d-Omega_3 | rs7970695 | 121423376 | A | G | -0.0253039 | 0.620549 | 12 | 0.00419603 | 1.20E-10 | 114999 | NA |
|  | met-d-Omega_3 | rs261290 | 58678720 | C | T | -0.114383 | 0.654653 | 15 | 0.00428189 | 3.90E-161 | 114999 | NA |
|  | met-d-Omega_3 | rs139974673 | 44027885 | C | T | 0.117987 | 0.025918 | 15 | 0.0128075 | 2.30E-21 | 114999 | NA |
|  | met-d-Omega_3 | rs34663616 | 58569330 | A | C | 0.0356728 | 0.137654 | 15 | 0.00602277 | 4.40E-10 | 114999 | NA |
|  | met-d-Omega_3 | rs633695 | 58725839 | G | A | 0.0840069 | 0.292348 | 15 | 0.00448287 | 9.10E-80 | 114999 | NA |
|  | met-d-Omega_3 | rs1672811 | 15501099 | C | T | 0.0251849 | 0.748488 | 16 | 0.0046967 | 3.00E-08 | 114999 | NA |
|  | met-d-Omega_3 | rs72789541 | 15127534 | A | T | -0.0810999 | 0.29597 | 16 | 0.00446068 | 5.60E-75 | 114999 | NA |
|  | met-d-Omega_3 | rs16940904 | 44186063 | T | C | -0.0355285 | 0.226571 | 17 | 0.0048772 | 3.90E-14 | 114999 | NA |
|  | met-d-Omega_3 | rs9304381 | 47158234 | T | C | 0.0528854 | 0.818434 | 18 | 0.00527814 | 5.20E-24 | 114999 | NA |
|  | met-d-Omega_3 | rs77960347 | 47109955 | G | A | 0.161749 | 0.013239 | 18 | 0.0177574 | 7.20E-22 | 114999 | NA |
|  | met-d-Omega_3 | rs737338 | 11347657 | T | C | -0.0726631 | 0.035186 | 19 | 0.0110354 | 3.50E-11 | 114999 | NA |
|  | met-d-Omega_3 | rs182611493 | 19458388 | G | A | -0.209571 | 0.012519 | 19 | 0.0195777 | 1.10E-27 | 114999 | NA |
|  | met-d-Omega_3 | rs58542926 | 19379549 | T | C | -0.171666 | 0.074383 | 19 | 0.00775231 | 1.40E-113 | 114999 | NA |
|  | met-d-Omega_3 | rs5112 | 45430280 | G | C | 0.0476852 | 0.533736 | 19 | 0.00437178 | 9.10E-30 | 114999 | NA |
|  | met-d-Omega_3 | rs1132899 | 45448036 | C | T | 0.0270834 | 0.509603 | 19 | 0.00410188 | 8.60E-11 | 114999 | NA |
|  | met-d-Omega_3 | rs157592 | 45424514 | C | A | 0.0284296 | 0.185267 | 19 | 0.00536368 | 3.60E-09 | 114999 | NA |
|  | met-d-Omega_3 | rs6129624 | 39167592 | A | G | -0.0257607 | 0.335237 | 20 | 0.00437946 | 5.10E-10 | 114999 | NA |
|  | met-d-Omega_3 | rs117143374 | 40555561 | C | T | -0.0370966 | 0.142254 | 21 | 0.005847 | 2.20E-10 | 114999 | NA |
|  | met-c-855 | rs1260326 | 27730940 | C | T | -0.096804 | 0.636884 | 2 | 0.012677 | 3.37E-14 | 13544 | 58.3114 |
|  | met-c-855 | rs174546 | 61569830 | T | C | -0.15381 | 0.40295 | 11 | 0.012443 | 1.19E-34 | 13544 | 152.7984 |
|  | met-c-855 | rs11604424 | 116651115 | T | C | -0.090079 | 0.756243 | 11 | 0.014237 | 3.32E-10 | 13540 | 40.0323 |
|  | met-c-855 | rs1077835 | 58723426 | G | A | 0.088821 | 0.249772 | 15 | 0.014466 | 1.08E-09 | 13538 | 37.6994 |
|  | met-c-855 | rs145717049 | 19130096 | T | C | -0.191179 | 0.044158 | 19 | 0.032739 | 6.67E-09 | 13536 | 34.0996 |
|  | met-c-855 | rs143988316 | 19667254 | T | C | -0.171397 | 0.069337 | 19 | 0.024385 | 2.95E-12 | 13539 | 49.4039 |
| Omega-3% | ebi-a-GCST90092932 | rs6693447 | 2330190 | G | T | 0.0255397 | 0.461701 | 1 | 0.00397732 | 1.40E-10 | 115006 | 41.2335 |
|  | ebi-a-GCST90092932 | rs638714 | 62906489 | T | G | -0.0322008 | 0.345965 | 1 | 0.00418219 | 1.40E-14 | 115006 | 59.2824 |
|  | ebi-a-GCST90092932 | rs11681659 | 136820960 | T | C | -0.0275604 | 0.716465 | 2 | 0.00437565 | 3.00E-10 | 115006 | 39.6722 |
|  | ebi-a-GCST90092932 | rs1260326 | 27730940 | C | T | -0.03861 | 0.604008 | 2 | 0.004051 | 1.60E-21 | 115006 | 90.8396 |
|  | ebi-a-GCST90092932 | rs6717316 | 79703384 | A | G | -0.0240496 | 0.670048 | 2 | 0.00420843 | 1.10E-08 | 115006 | 32.6569 |
|  | ebi-a-GCST90092932 | rs4860987 | 69491284 | T | A | 0.0394847 | 0.258612 | 4 | 0.00478848 | 1.60E-16 | 115006 | 67.9927 |
|  | ebi-a-GCST90092932 | rs11379773 | 87695669 | GT | G | 0.0274232 | 0.213412 | 5 | 0.00484826 | 1.50E-08 | 115006 | 31.9937 |
|  | ebi-a-GCST90092932 | rs272888 | 131665423 | C | T | 0.028298 | 0.706966 | 5 | 0.00434971 | 7.70E-11 | 115006 | 42.3244 |
|  | ebi-a-GCST90092932 | rs662138 | 160564476 | G | C | -0.0312278 | 0.185912 | 6 | 0.0050918 | 8.60E-10 | 115006 | 37.6132 |
|  | ebi-a-GCST90092932 | rs4000713 | 25990597 | A | G | -0.0267482 | 0.295416 | 7 | 0.00434956 | 7.80E-10 | 115006 | 37.8180 |
|  | ebi-a-GCST90092932 | rs62466318 | 73042085 | T | C | -0.0404238 | 0.204201 | 7 | 0.00493739 | 2.70E-16 | 115006 | 67.0316 |
|  | ebi-a-GCST90092932 | rs112875651 | 126506694 | A | G | -0.0517835 | 0.392337 | 8 | 0.00411717 | 2.80E-36 | 115006 | 158.1922 |
|  | ebi-a-GCST90092932 | rs7924036 | 65191645 | T | G | 0.0374032 | 0.504218 | 10 | 0.00396292 | 3.80E-21 | 115006 | 89.0814 |
|  | ebi-a-GCST90092932 | rs61886804 | 96749936 | G | T | 0.0351105 | 0.200238 | 10 | 0.0049535 | 1.40E-12 | 115006 | 50.2400 |
|  | ebi-a-GCST90092932 | rs77323894 | 60896505 | C | G | 0.0673413 | 0.038434 | 11 | 0.0107043 | 3.20E-10 | 115006 | 39.5773 |
|  | ebi-a-GCST90092932 | rs964184 | 116648917 | C | G | -0.0388695 | 0.867233 | 11 | 0.00580799 | 2.20E-11 | 115006 | 44.7885 |
|  | ebi-a-GCST90092932 | rs150370599 | 60718792 | T | C | 0.0472803 | 0.080318 | 11 | 0.00726173 | 7.50E-11 | 115006 | 42.3916 |
|  | ebi-a-GCST90092932 | rs12226389 | 61823630 | C | T | -0.0642431 | 0.185805 | 11 | 0.00511013 | 3.00E-36 | 115006 | 158.0480 |
|  | ebi-a-GCST90092932 | rs508049 | 68675497 | T | C | -0.10589 | 0.049239 | 11 | 0.009204 | 1.20E-30 | 115006 | 132.3600 |
|  | ebi-a-GCST90092932 | rs195445 | 61744342 | T | C | -0.038133 | 0.688414 | 11 | 0.00426447 | 3.80E-19 | 115006 | 79.9598 |
|  | ebi-a-GCST90092932 | rs7944950 | 75505827 | C | T | -0.0741144 | 0.085668 | 11 | 0.00706143 | 9.00E-26 | 115006 | 110.1590 |
|  | ebi-a-GCST90092932 | rs145786300 | 61406089 | A | G | -0.188873 | 0.011993 | 11 | 0.0187215 | 6.20E-24 | 115006 | 101.7791 |
|  | ebi-a-GCST90092932 | rs174564 | 61588305 | G | A | -0.391303 | 0.347009 | 11 | 0.00412995 | 1.00E-200 | 115006 | 8977.1146 |
|  | ebi-a-GCST90092932 | rs9563335 | 56069705 | G | A | -0.0444873 | 0.861572 | 13 | 0.00771128 | 8.00E-09 | 115006 | 33.2827 |
|  | ebi-a-GCST90092932 | rs1532085 | 58683366 | G | A | -0.078198 | 0.613968 | 15 | 0.0040709 | 3.10E-82 | 115006 | 368.9865 |
|  | ebi-a-GCST90092932 | rs1077835 | 58723426 | G | A | 0.0910667 | 0.219711 | 15 | 0.00479257 | 1.70E-80 | 115006 | 361.0624 |
|  | ebi-a-GCST90092932 | rs139974673 | 44027885 | C | T | 0.083441 | 0.025916 | 15 | 0.0124929 | 2.40E-11 | 115006 | 44.6100 |
|  | ebi-a-GCST90092932 | rs72789541 | 15127534 | A | T | -0.0992036 | 0.295973 | 16 | 0.00434806 | 3.20E-115 | 115006 | 520.5516 |
|  | ebi-a-GCST90092932 | rs35390787 | 15678414 | CA | C | 0.0273295 | 0.438254 | 16 | 0.00408187 | 2.20E-11 | 115006 | 44.8276 |
|  | ebi-a-GCST90092932 | rs78423941 | 15421686 | C | G | 0.0268058 | 0.396267 | 16 | 0.00486895 | 3.70E-08 | 115006 | 30.3101 |
|  | ebi-a-GCST90092932 | rs8074191 | 17407191 | C | T | -0.0268897 | 0.756121 | 17 | 0.00462305 | 6.00E-09 | 115006 | 33.8310 |
|  | ebi-a-GCST90092932 | rs16940904 | 44186063 | T | C | -0.041183 | 0.226566 | 17 | 0.00475547 | 4.70E-18 | 115006 | 74.9979 |
|  | ebi-a-GCST90092932 | rs9947684 | 47166694 | G | A | 0.0282504 | 0.654466 | 18 | 0.00416339 | 1.20E-11 | 115006 | 46.0421 |
|  | ebi-a-GCST90092932 | rs182611493 | 19458388 | G | A | -0.181074 | 0.012519 | 19 | 0.0190938 | 2.50E-21 | 115006 | 89.9347 |
|  | ebi-a-GCST90092932 | rs190921611 | 45441475 | A | G | 0.0314663 | 0.311083 | 19 | 0.00462576 | 1.00E-11 | 115006 | 46.2727 |
|  | ebi-a-GCST90092932 | rs58542926 | 19379549 | T | C | -0.132116 | 0.074379 | 19 | 0.00756045 | 2.20E-68 | 115006 | 305.3624 |
|  | ebi-a-GCST90092932 | rs72654473 | 45414399 | A | C | 0.0390547 | 0.108331 | 19 | 0.00639034 | 9.90E-10 | 115006 | 37.3507 |
|  | met-d-Omega_3_pct | rs638714 | 62906489 | T | G | -0.0328872 | 0.345963 | 1 | 0.00428078 | 4.90E-15 | 114999 | NA |
|  | met-d-Omega_3_pct | rs6693447 | 2330190 | G | T | 0.0263349 | 0.461686 | 1 | 0.00407105 | 8.30E-12 | 114999 | NA |
|  | met-d-Omega_3_pct | rs2011946 | 136817616 | A | C | -0.0269984 | 0.734729 | 2 | 0.00458772 | 2.60E-09 | 114999 | NA |
|  | met-d-Omega_3_pct | rs1260326 | 27730940 | C | T | -0.0376379 | 0.60401 | 2 | 0.00414705 | 2.30E-19 | 114999 | NA |
|  | met-d-Omega_3_pct | rs4860987 | 69491284 | T | A | 0.0407766 | 0.258609 | 4 | 0.00490097 | 8.10E-17 | 114999 | NA |
|  | met-d-Omega_3_pct | rs272888 | 131665423 | C | T | 0.0285458 | 0.706952 | 5 | 0.00445249 | 1.80E-11 | 114999 | NA |
|  | met-d-Omega_3_pct | rs7444298 | 87730027 | G | A | 0.0268334 | 0.236979 | 5 | 0.00477252 | 2.40E-08 | 114999 | NA |
|  | met-d-Omega_3_pct | rs2394976 | 31311912 | T | G | -0.0312159 | 0.161648 | 6 | 0.00550023 | 5.40E-09 | 114999 | NA |
|  | met-d-Omega_3_pct | rs662138 | 160564476 | G | C | -0.0338037 | 0.18591 | 6 | 0.00521203 | 6.60E-11 | 114999 | NA |
|  | met-d-Omega_3_pct | rs4000713 | 25990597 | A | G | -0.0273379 | 0.295408 | 7 | 0.00445218 | 1.60E-10 | 114999 | NA |
|  | met-d-Omega_3_pct | rs62466318 | 73042085 | T | C | -0.0409148 | 0.204178 | 7 | 0.00505439 | 3.10E-16 | 114999 | NA |
|  | met-d-Omega_3_pct | rs73109460 | 44785800 | A | G | -0.0320063 | 0.123622 | 7 | 0.00620457 | 7.80E-09 | 114999 | NA |
|  | met-d-Omega_3_pct | rs112875651 | 126506694 | A | G | -0.0517841 | 0.392346 | 8 | 0.00421392 | 5.70E-35 | 114999 | NA |
|  | met-d-Omega_3_pct | rs2236514 | 139572068 | G | C | -0.0222949 | 0.657511 | 9 | 0.00433047 | 3.10E-08 | 114999 | NA |
|  | met-d-Omega_3_pct | rs7924036 | 65191645 | T | G | 0.0384969 | 0.504205 | 10 | 0.00405633 | 4.00E-23 | 114999 | NA |
|  | met-d-Omega_3_pct | rs56233220 | 96717286 | C | G | 0.0358845 | 0.200191 | 10 | 0.00507602 | 4.40E-13 | 114999 | NA |
|  | met-d-Omega_3_pct | rs11236512 | 75449819 | A | C | -0.0713676 | 0.062661 | 11 | 0.00927047 | 5.30E-15 | 114999 | NA |
|  | met-d-Omega_3_pct | rs2232143 | 60899701 | C | T | 0.107875 | 0.021879 | 11 | 0.0144553 | 1.30E-13 | 114999 | NA |
|  | met-d-Omega_3_pct | rs191623731 | 61844663 | G | T | 0.118026 | 0.01584 | 11 | 0.0161995 | 9.80E-14 | 114999 | NA |
|  | met-d-Omega_3_pct | rs964184 | 116648917 | C | G | -0.0379163 | 0.867229 | 11 | 0.00594666 | 1.20E-10 | 114999 | NA |
|  | met-d-Omega_3_pct | rs143355652 | 61453822 | T | C | -0.179283 | 0.010467 | 11 | 0.0203444 | 1.20E-18 | 114999 | NA |
|  | met-d-Omega_3_pct | rs174564 | 61588305 | G | A | -0.391514 | 0.347013 | 11 | 0.00422879 | 1.00E-200 | 114999 | NA |
|  | met-d-Omega_3_pct | rs11230829 | 61701898 | G | A | -0.11592 | 0.027786 | 11 | 0.0147756 | 1.70E-15 | 114999 | NA |
|  | met-d-Omega_3_pct | rs75227397 | 75453974 | A | G | -0.0631164 | 0.031062 | 11 | 0.0117901 | 2.80E-08 | 114999 | NA |
|  | met-d-Omega_3_pct | rs12226389 | 61823630 | C | T | -0.0616476 | 0.18582 | 11 | 0.00523223 | 2.20E-32 | 114999 | NA |
|  | met-d-Omega_3_pct | rs149402055 | 68525539 | T | C | -0.173399 | 0.019709 | 11 | 0.015229 | 9.20E-30 | 114999 | NA |
|  | met-d-Omega_3_pct | rs145786300 | 61406089 | A | G | -0.186883 | 0.011985 | 11 | 0.019176 | 1.30E-22 | 114999 | NA |
|  | met-d-Omega_3_pct | rs9563335 | 56069705 | G | A | -0.0442419 | 0.861577 | 13 | 0.00789332 | 5.50E-09 | 114999 | NA |
|  | met-d-Omega_3_pct | rs261291 | 58680178 | C | T | 0.0807217 | 0.356068 | 15 | 0.00424237 | 1.80E-83 | 114999 | NA |
|  | met-d-Omega_3_pct | rs11632618 | 58724706 | A | G | 0.0776424 | 0.069927 | 15 | 0.0079442 | 1.10E-22 | 114999 | NA |
|  | met-d-Omega_3_pct | rs1560390 | 58580781 | C | T | -0.0324585 | 0.219684 | 15 | 0.00492881 | 6.90E-12 | 114999 | NA |
|  | met-d-Omega_3_pct | rs139974673 | 44027885 | C | T | 0.0839134 | 0.025918 | 15 | 0.0127875 | 1.30E-11 | 114999 | NA |
|  | met-d-Omega_3_pct | rs35390787 | 15678414 | C | CA | -0.0268978 | 0.561747 | 16 | 0.00417832 | 5.60E-11 | 114999 | NA |
|  | met-d-Omega_3_pct | rs72789541 | 15127534 | A | T | -0.0970051 | 0.29597 | 16 | 0.0044509 | 3.50E-106 | 114999 | NA |
|  | met-d-Omega_3_pct | rs16940904 | 44186063 | T | C | -0.0421552 | 0.226571 | 17 | 0.00486779 | 2.40E-19 | 114999 | NA |
|  | met-d-Omega_3_pct | rs8074191 | 17407191 | C | T | -0.0265115 | 0.756127 | 17 | 0.00473237 | 2.30E-08 | 114999 | NA |
|  | met-d-Omega_3_pct | rs9947684 | 47166694 | G | A | 0.0289177 | 0.654493 | 18 | 0.00426163 | 2.80E-12 | 114999 | NA |
|  | met-d-Omega_3_pct | rs58542926 | 19379549 | T | C | -0.131159 | 0.074383 | 19 | 0.00773888 | 1.20E-67 | 114999 | NA |
|  | met-d-Omega_3_pct | rs72654473 | 45414399 | A | C | 0.0365241 | 0.108333 | 19 | 0.00654126 | 6.10E-09 | 114999 | NA |
|  | met-d-Omega_3_pct | rs182611493 | 19458388 | G | A | -0.172066 | 0.012519 | 19 | 0.0195438 | 8.90E-19 | 114999 | NA |
|  | met-d-Omega_3_pct | rs190921611 | 45441475 | A | G | 0.031631 | 0.311094 | 19 | 0.00473503 | 2.00E-11 | 114999 | NA |
| Omega-6 | ebi-a-GCST90092933 | rs534417 | 23784965 | G | A | 0.0391707 | 0.875008 | 1 | 0.00607383 | 1.10E-10 | 115006 | 41.5908 |
|  | ebi-a-GCST90092933 | rs2986164 | 25622291 | A | G | -0.0247791 | 0.535868 | 1 | 0.00440639 | 1.90E-08 | 115006 | 31.6232 |
|  | ebi-a-GCST90092933 | rs34232196 | 55489542 | T | C | -0.0327764 | 0.245336 | 1 | 0.00471815 | 3.70E-12 | 115006 | 48.2591 |
|  | ebi-a-GCST90092933 | rs1002687 | 62963737 | A | G | 0.0917563 | 0.644739 | 1 | 0.0042098 | 2.50E-105 | 115006 | 475.0604 |
|  | ebi-a-GCST90092933 | rs602633 | 109821511 | G | T | 0.0573983 | 0.782937 | 1 | 0.00487783 | 5.80E-32 | 115006 | 138.4665 |
|  | ebi-a-GCST90092933 | rs496654 | 234851165 | C | A | 0.0302882 | 0.517238 | 1 | 0.00403175 | 5.80E-14 | 115006 | 56.4365 |
|  | ebi-a-GCST90092933 | rs870526 | 20369562 | T | C | -0.0320783 | 0.521425 | 2 | 0.00403664 | 1.90E-15 | 115006 | 63.1514 |
|  | ebi-a-GCST90092933 | rs3770586 | 169828995 | T | C | -0.0229768 | 0.48398 | 2 | 0.00404862 | 1.40E-08 | 115006 | 32.2081 |
|  | ebi-a-GCST90092933 | rs1260326 | 27730940 | C | T | -0.064724 | 0.604008 | 2 | 0.00411907 | 1.20E-55 | 115006 | 246.9064 |
|  | ebi-a-GCST90092933 | rs672889 | 21319016 | G | T | 0.0758525 | 0.860074 | 2 | 0.00580072 | 4.50E-39 | 115006 | 170.9921 |
|  | ebi-a-GCST90092933 | rs6547409 | 21190209 | T | C | -0.0808582 | 0.051259 | 2 | 0.00924119 | 2.10E-18 | 115006 | 76.5583 |
|  | ebi-a-GCST90092933 | rs4299376 | 44072576 | T | G | -0.0356946 | 0.676282 | 2 | 0.00431655 | 1.30E-16 | 115006 | 68.3804 |
|  | ebi-a-GCST90092933 | rs4860948 | 69340991 | A | T | 0.0285942 | 0.244351 | 4 | 0.00470392 | 1.20E-09 | 115006 | 36.9518 |
|  | ebi-a-GCST90092933 | rs13108218 | 3443931 | G | A | -0.035128 | 0.615408 | 4 | 0.00417284 | 3.80E-17 | 115006 | 70.8669 |
|  | ebi-a-GCST90092933 | rs4704210 | 74635225 | C | G | 0.0463297 | 0.374206 | 5 | 0.00416513 | 9.70E-29 | 115006 | 123.7263 |
|  | ebi-a-GCST90092933 | rs7707394 | 74472939 | A | G | 0.0294421 | 0.357102 | 5 | 0.00419959 | 2.40E-12 | 115006 | 49.1500 |
|  | ebi-a-GCST90092933 | rs6882345 | 156397673 | A | G | 0.0441725 | 0.63285 | 5 | 0.00417626 | 3.80E-26 | 115006 | 111.8739 |
|  | ebi-a-GCST90092933 | rs28377109 | 32434716 | A | C | 0.0450439 | 0.116077 | 6 | 0.00629445 | 8.30E-13 | 115006 | 51.2102 |
|  | ebi-a-GCST90092933 | rs9266229 | 31325323 | G | C | 0.027057 | 0.433975 | 6 | 0.00413903 | 6.30E-11 | 115006 | 42.7329 |
|  | ebi-a-GCST90092933 | rs7750288 | 160400147 | G | A | 0.025103 | 0.285016 | 6 | 0.00445908 | 1.80E-08 | 115006 | 31.6928 |
|  | ebi-a-GCST90092933 | rs6938647 | 160986915 | C | A | -0.0478033 | 0.782332 | 6 | 0.00494492 | 4.20E-22 | 115006 | 93.4539 |
|  | ebi-a-GCST90092933 | rs9295128 | 160751531 | T | G | -0.19728 | 0.016832 | 6 | 0.0159431 | 3.60E-35 | 115006 | 153.1160 |
|  | ebi-a-GCST90092933 | rs11755689 | 32586376 | G | A | 0.0416405 | 0.334092 | 6 | 0.00440473 | 3.30E-21 | 115006 | 89.3703 |
|  | ebi-a-GCST90092933 | rs3756772 | 116325142 | T | C | 0.0232823 | 0.400766 | 6 | 0.00410898 | 1.50E-08 | 115006 | 32.1058 |
|  | ebi-a-GCST90092933 | rs55747707 | 73037366 | A | G | -0.0498035 | 0.203638 | 7 | 0.00501986 | 3.40E-23 | 115006 | 98.4320 |
|  | ebi-a-GCST90092933 | rs2126259 | 9185146 | C | T | 0.0829891 | 0.899179 | 8 | 0.00669742 | 2.90E-35 | 115006 | 153.5419 |
|  | ebi-a-GCST90092933 | rs6471717 | 59377357 | A | G | -0.0295948 | 0.663076 | 8 | 0.00427438 | 4.40E-12 | 115006 | 47.9385 |
|  | ebi-a-GCST90092933 | rs112875651 | 126506694 | A | G | -0.0635591 | 0.392337 | 8 | 0.00418539 | 4.40E-52 | 115006 | 230.6129 |
|  | ebi-a-GCST90092933 | rs2721961 | 116657911 | G | T | -0.0274809 | 0.280836 | 8 | 0.00449395 | 9.70E-10 | 115006 | 37.3943 |
|  | ebi-a-GCST90092933 | rs7831074 | 141633257 | G | C | 0.0276942 | 0.759123 | 8 | 0.00504475 | 4.00E-08 | 115006 | 30.1369 |
|  | ebi-a-GCST90092933 | rs11789603 | 107647019 | T | C | 0.0483698 | 0.108847 | 9 | 0.00648531 | 8.80E-14 | 115006 | 55.6272 |
|  | ebi-a-GCST90092933 | rs2740488 | 107661742 | C | A | -0.0496352 | 0.265318 | 9 | 0.00457594 | 2.10E-27 | 115006 | 117.6573 |
|  | ebi-a-GCST90092933 | rs4008004 | 15300968 | A | C | 0.0334374 | 0.221833 | 9 | 0.00487192 | 6.70E-12 | 115006 | 47.1048 |
|  | ebi-a-GCST90092933 | rs115478735 | 136149711 | T | A | 0.0424072 | 0.183303 | 9 | 0.00521297 | 4.10E-16 | 115006 | 66.1773 |
|  | ebi-a-GCST90092933 | rs148063610 | 45998984 | CAAATAAAT | C | 0.0320419 | 0.236598 | 10 | 0.00484243 | 3.70E-11 | 115006 | 43.7834 |
|  | ebi-a-GCST90092933 | rs117488242 | 5261832 | G | A | -0.0355756 | 0.131672 | 10 | 0.00624596 | 1.20E-08 | 115006 | 32.4419 |
|  | ebi-a-GCST90092933 | rs3817335 | 47643891 | A | T | -0.027965 | 0.35055 | 11 | 0.00421165 | 3.10E-11 | 115006 | 44.0885 |
|  | ebi-a-GCST90092933 | rs964184 | 116648917 | C | G | -0.138873 | 0.867233 | 11 | 0.00594037 | 7.19E-121 | 115006 | 546.5233 |
|  | ebi-a-GCST90092933 | rs72997616 | 75474195 | A | C | -0.0527256 | 0.094062 | 11 | 0.00693794 | 3.00E-14 | 115006 | 57.7540 |
|  | ebi-a-GCST90092933 | rs200671503 | 116817978 | TTA | T | -0.0501356 | 0.058035 | 11 | 0.00866302 | 7.20E-09 | 115006 | 33.4930 |
|  | ebi-a-GCST90092933 | rs35350651 | 111907431 | AC | A | 0.0277979 | 0.502598 | 12 | 0.00403844 | 5.80E-12 | 115006 | 47.3802 |
|  | ebi-a-GCST90092933 | rs7139079 | 121415293 | A | G | -0.0292329 | 0.592509 | 12 | 0.00411236 | 1.20E-12 | 115006 | 50.5314 |
|  | ebi-a-GCST90092933 | rs6602911 | 114547372 | T | C | 0.0262676 | 0.360074 | 13 | 0.00419748 | 3.90E-10 | 115006 | 39.1619 |
|  | ebi-a-GCST90092933 | rs261290 | 58678720 | C | T | -0.0965237 | 0.654648 | 15 | 0.00424783 | 2.70E-114 | 115006 | 516.3375 |
|  | ebi-a-GCST90092933 | rs633695 | 58725839 | G | A | 0.0730669 | 0.292334 | 15 | 0.00444717 | 1.20E-60 | 115006 | 269.9441 |
|  | ebi-a-GCST90092933 | rs3764261 | 56993324 | A | C | 0.0616184 | 0.324448 | 16 | 0.00430576 | 1.90E-46 | 115006 | 204.7959 |
|  | ebi-a-GCST90092933 | rs76116020 | 69385641 | G | A | -0.0570097 | 0.043763 | 16 | 0.00986958 | 7.60E-09 | 115006 | 33.3657 |
|  | ebi-a-GCST90092933 | rs56325564 | 45766771 | A | G | 0.0227535 | 0.482656 | 17 | 0.00406151 | 2.10E-08 | 115006 | 31.3849 |
|  | ebi-a-GCST90092933 | rs740516 | 67082962 | G | C | -0.0313763 | 0.151132 | 17 | 0.00565493 | 2.90E-08 | 115006 | 30.7857 |
|  | ebi-a-GCST90092933 | rs77960347 | 47109955 | G | A | 0.272485 | 0.013234 | 18 | 0.0176065 | 5.00E-54 | 115006 | 239.5185 |
|  | ebi-a-GCST90092933 | rs9304381 | 47158234 | T | C | 0.0700213 | 0.818428 | 18 | 0.0052323 | 7.70E-41 | 115006 | 179.0915 |
|  | ebi-a-GCST90092933 | rs58542926 | 19379549 | T | C | -0.1284 | 0.074379 | 19 | 0.00768267 | 1.10E-62 | 115006 | 279.3224 |
|  | ebi-a-GCST90092933 | rs56322906 | 11346155 | A | G | -0.101934 | 0.035155 | 19 | 0.0109515 | 1.30E-20 | 115006 | 86.6345 |
|  | ebi-a-GCST90092933 | rs74747585 | 45039971 | C | T | -0.0737645 | 0.025172 | 19 | 0.0132168 | 2.40E-08 | 115006 | 31.1489 |
|  | ebi-a-GCST90092933 | rs1081105 | 45412955 | C | A | 0.118624 | 0.027622 | 19 | 0.0123232 | 6.20E-22 | 115006 | 92.6612 |
|  | ebi-a-GCST90092933 | rs142158911 | 11190534 | A | G | -0.0937617 | 0.116703 | 19 | 0.00631307 | 6.80E-50 | 115006 | 220.5818 |
|  | ebi-a-GCST90092933 | rs79429216 | 45445517 | A | G | 0.151678 | 0.012712 | 19 | 0.0179494 | 2.90E-17 | 115006 | 71.4077 |
|  | ebi-a-GCST90092933 | rs1065853 | 45413233 | T | G | -0.198416 | 0.080578 | 19 | 0.00743205 | 5.11E-157 | 115006 | 712.7482 |
|  | ebi-a-GCST90092933 | rs111278137 | 45215081 | A | G | -0.0787637 | 0.021615 | 19 | 0.0143417 | 4.00E-08 | 115006 | 30.1614 |
|  | ebi-a-GCST90092933 | rs2378390 | 34150207 | A | G | -0.0332858 | 0.140867 | 20 | 0.0058156 | 1.00E-08 | 115006 | 32.7589 |
|  | ebi-a-GCST90092933 | rs1883711 | 39179822 | C | G | 0.0905557 | 0.031197 | 20 | 0.0118431 | 2.10E-14 | 115006 | 58.4657 |
|  | ebi-a-GCST90092933 | rs1800961 | 43042364 | T | C | -0.0748822 | 0.030194 | 20 | 0.0117724 | 2.00E-10 | 115006 | 40.4601 |
|  | ebi-a-GCST90092933 | rs5754102 | 21916272 | A | C | -0.0326814 | 0.183238 | 22 | 0.00527073 | 5.60E-10 | 115006 | 38.4468 |
|  | ebi-a-GCST90092933 | rs9616847 | 50868669 | T | A | 0.0235066 | 0.388308 | 22 | 0.00415553 | 1.50E-08 | 115006 | 31.9983 |
|  | met-d-Omega_6 | rs199900492 | 234850420 | CA | C | -0.0306276 | 0.484009 | 1 | 0.00404392 | 2.30E-15 | 114999 | NA |
|  | met-d-Omega_6 | rs534417 | 23784965 | G | A | 0.0390397 | 0.875005 | 1 | 0.00607823 | 9.30E-11 | 114999 | NA |
|  | met-d-Omega_6 | rs200730299 | 55491853 | C | A | -0.0365309 | 0.194476 | 1 | 0.00542087 | 7.10E-12 | 114999 | NA |
|  | met-d-Omega_6 | rs2986164 | 25622291 | A | G | -0.0250071 | 0.535856 | 1 | 0.00440965 | 3.40E-09 | 114999 | NA |
|  | met-d-Omega_6 | rs12740374 | 109817590 | T | G | -0.0572469 | 0.221071 | 1 | 0.00485222 | 1.50E-32 | 114999 | NA |
|  | met-d-Omega_6 | rs1002687 | 62963737 | A | G | 0.090971 | 0.644748 | 1 | 0.00421287 | 1.00E-107 | 114999 | NA |
|  | met-d-Omega_6 | rs1260326 | 27730940 | C | T | -0.0642561 | 0.60401 | 2 | 0.00412206 | 3.90E-55 | 114999 | NA |
|  | met-d-Omega_6 | rs870526 | 20369562 | T | C | -0.0318914 | 0.521426 | 2 | 0.00403955 | 7.70E-16 | 114999 | NA |
|  | met-d-Omega_6 | rs4299376 | 44072576 | T | G | -0.0353905 | 0.676279 | 2 | 0.00431976 | 1.10E-16 | 114999 | NA |
|  | met-d-Omega_6 | rs6547409 | 21190209 | T | C | -0.0813514 | 0.051258 | 2 | 0.00924809 | 2.40E-20 | 114999 | NA |
|  | met-d-Omega_6 | rs672889 | 21319016 | G | T | 0.0763943 | 0.860091 | 2 | 0.00580512 | 1.30E-41 | 114999 | NA |
|  | met-d-Omega_6 | rs3770586 | 169828995 | T | C | -0.0229674 | 0.48397 | 2 | 0.00405156 | 7.10E-09 | 114999 | NA |
|  | met-d-Omega_6 | rs13108218 | 3443931 | G | A | -0.035165 | 0.615417 | 4 | 0.00417574 | 3.60E-18 | 114999 | NA |
|  | met-d-Omega_6 | rs4860948 | 69340991 | A | T | 0.027907 | 0.244353 | 4 | 0.00470731 | 1.90E-09 | 114999 | NA |
|  | met-d-Omega_6 | rs4704210 | 74635225 | C | G | 0.0467287 | 0.374212 | 5 | 0.00416788 | 6.10E-30 | 114999 | NA |
|  | met-d-Omega_6 | rs6882345 | 156397673 | A | G | 0.0448784 | 0.632863 | 5 | 0.00417905 | 1.20E-27 | 114999 | NA |
|  | met-d-Omega_6 | rs7707394 | 74472939 | A | G | 0.02951 | 0.357106 | 5 | 0.00420247 | 1.20E-12 | 114999 | NA |
|  | met-d-Omega_6 | rs6934962 | 116322349 | T | C | 0.0228258 | 0.400489 | 6 | 0.00411241 | 2.20E-08 | 114999 | NA |
|  | met-d-Omega_6 | rs3734854 | 31078836 | A | G | 0.0477116 | 0.064679 | 6 | 0.0082133 | 5.90E-11 | 114999 | NA |
|  | met-d-Omega_6 | rs35603463 | 32531745 | C | T | 0.0336386 | 0.566969 | 6 | 0.00484067 | 4.60E-10 | 114999 | NA |
|  | met-d-Omega_6 | rs7750288 | 160400147 | G | A | 0.0249212 | 0.285011 | 6 | 0.00446247 | 1.30E-08 | 114999 | NA |
|  | met-d-Omega_6 | rs9295128 | 160751531 | T | G | -0.196375 | 0.016833 | 6 | 0.0159543 | 3.40E-36 | 114999 | NA |
|  | met-d-Omega_6 | rs114863007 | 34729158 | A | G | -0.0457294 | 0.094543 | 6 | 0.00693029 | 7.30E-12 | 114999 | NA |
|  | met-d-Omega_6 | rs28383314 | 32587213 | C | T | 0.0388549 | 0.622939 | 6 | 0.00415994 | 1.70E-18 | 114999 | NA |
|  | met-d-Omega_6 | rs6938647 | 160986915 | C | A | -0.0481657 | 0.782336 | 6 | 0.00494853 | 1.90E-23 | 114999 | NA |
|  | met-d-Omega_6 | rs55747707 | 73037366 | A | G | -0.0494876 | 0.203615 | 7 | 0.0050239 | 1.70E-22 | 114999 | NA |
|  | met-d-Omega_6 | rs6471717 | 59377357 | A | G | -0.0290285 | 0.663091 | 8 | 0.00427754 | 4.00E-12 | 114999 | NA |
|  | met-d-Omega_6 | rs112875651 | 126506694 | A | G | -0.063694 | 0.392346 | 8 | 0.00418831 | 2.20E-53 | 114999 | NA |
|  | met-d-Omega_6 | rs2737245 | 116658583 | T | G | -0.0274764 | 0.278598 | 8 | 0.00450795 | 1.40E-09 | 114999 | NA |
|  | met-d-Omega_6 | rs1461729 | 9187242 | G | A | 0.0835465 | 0.899221 | 8 | 0.00670616 | 2.80E-36 | 114999 | NA |
|  | met-d-Omega_6 | rs7831074 | 141633257 | G | C | 0.0278428 | 0.759127 | 8 | 0.00504852 | 4.60E-08 | 114999 | NA |
|  | met-d-Omega_6 | rs4008004 | 15300968 | A | C | 0.0329594 | 0.221842 | 9 | 0.00487542 | 8.40E-12 | 114999 | NA |
|  | met-d-Omega_6 | rs11789603 | 107647019 | T | C | 0.0478798 | 0.108831 | 9 | 0.00649064 | 9.70E-14 | 114999 | NA |
|  | met-d-Omega_6 | rs2740488 | 107661742 | C | A | -0.0501345 | 0.265321 | 9 | 0.00457926 | 5.40E-28 | 114999 | NA |
|  | met-d-Omega_6 | rs115478735 | 136149711 | T | A | 0.0422787 | 0.183315 | 9 | 0.00521674 | 2.20E-17 | 114999 | NA |
|  | met-d-Omega_6 | rs75406471 | 5257647 | A | G | -0.0314077 | 0.154684 | 10 | 0.00559075 | 2.70E-08 | 114999 | NA |
|  | met-d-Omega_6 | rs148063610 | 45998984 | C | CAAATAAAT | -0.0316205 | 0.763414 | 10 | 0.00484601 | 4.00E-11 | 114999 | NA |
|  | met-d-Omega_6 | rs141469619 | 116714293 | G | A | 0.110551 | 0.01014 | 11 | 0.0213113 | 1.40E-08 | 114999 | NA |
|  | met-d-Omega_6 | rs964184 | 116648917 | C | G | -0.138896 | 0.867229 | 11 | 0.00594434 | 1.10E-125 | 114999 | NA |
|  | met-d-Omega_6 | rs72997616 | 75474195 | A | C | -0.051564 | 0.094064 | 11 | 0.00694288 | 1.60E-13 | 114999 | NA |
|  | met-d-Omega_6 | rs3817335 | 47643891 | A | T | -0.0277621 | 0.350559 | 11 | 0.00421464 | 9.80E-12 | 114999 | NA |
|  | met-d-Omega_6 | rs200671503 | 116817978 | T | TTA | 0.0508862 | 0.941966 | 11 | 0.00866917 | 2.10E-09 | 114999 | NA |
|  | met-d-Omega_6 | rs4766578 | 111904371 | A | T | 0.0277794 | 0.503267 | 12 | 0.00404125 | 1.50E-12 | 114999 | NA |
|  | met-d-Omega_6 | rs7139079 | 121415293 | A | G | -0.0297379 | 0.592501 | 12 | 0.00411541 | 3.30E-13 | 114999 | NA |
|  | met-d-Omega_6 | rs6602911 | 114547372 | T | C | 0.0257493 | 0.360079 | 13 | 0.00420044 | 1.30E-09 | 114999 | NA |
|  | met-d-Omega_6 | rs633695 | 58725839 | G | A | 0.0725178 | 0.292348 | 15 | 0.00445048 | 1.30E-59 | 114999 | NA |
|  | met-d-Omega_6 | rs261290 | 58678720 | C | T | -0.0965507 | 0.654653 | 15 | 0.00425095 | 1.00E-116 | 114999 | NA |
|  | met-d-Omega_6 | rs183130 | 56991363 | T | C | 0.0615133 | 0.324072 | 16 | 0.00431123 | 1.40E-48 | 114999 | NA |
|  | met-d-Omega_6 | rs740516 | 67082962 | G | C | -0.0315718 | 0.151132 | 17 | 0.00565892 | 1.40E-08 | 114999 | NA |
|  | met-d-Omega_6 | rs4439799 | 45781599 | T | C | 0.0224228 | 0.502274 | 17 | 0.00404559 | 1.30E-08 | 114999 | NA |
|  | met-d-Omega_6 | rs77960347 | 47109955 | G | A | 0.275526 | 0.013239 | 18 | 0.0176159 | 2.80E-56 | 114999 | NA |
|  | met-d-Omega_6 | rs9304381 | 47158234 | T | C | 0.0702336 | 0.818434 | 18 | 0.00523609 | 7.20E-42 | 114999 | NA |
|  | met-d-Omega_6 | rs56322906 | 11346155 | A | G | -0.100371 | 0.035157 | 19 | 0.0109592 | 1.20E-19 | 114999 | NA |
|  | met-d-Omega_6 | rs142158911 | 11190534 | A | G | -0.0944019 | 0.116705 | 19 | 0.00631781 | 5.20E-52 | 114999 | NA |
|  | met-d-Omega_6 | rs58542926 | 19379549 | T | C | -0.128057 | 0.074383 | 19 | 0.0076883 | 2.50E-65 | 114999 | NA |
|  | met-d-Omega_6 | rs1081105 | 45412955 | C | A | 0.118778 | 0.027624 | 19 | 0.0123322 | 1.80E-22 | 114999 | NA |
|  | met-d-Omega_6 | rs79429216 | 45445517 | A | G | 0.15083 | 0.012713 | 19 | 0.0179625 | 1.30E-17 | 114999 | NA |
|  | met-d-Omega_6 | rs1065853 | 45413233 | T | G | -0.198623 | 0.080578 | 19 | 0.00743767 | 3.60E-160 | 114999 | NA |
|  | met-d-Omega_6 | rs1883711 | 39179822 | C | G | 0.0922513 | 0.031191 | 20 | 0.0118528 | 3.20E-16 | 114999 | NA |
|  | met-d-Omega_6 | rs2378390 | 34150207 | A | G | -0.0333321 | 0.140871 | 20 | 0.00581966 | 3.20E-09 | 114999 | NA |
|  | met-d-Omega_6 | rs1800961 | 43042364 | T | C | -0.0744567 | 0.030192 | 20 | 0.0117812 | 3.30E-10 | 114999 | NA |
|  | met-d-Omega_6 | rs5754102 | 21916272 | A | C | -0.0316 | 0.183245 | 22 | 0.00527441 | 9.90E-10 | 114999 | NA |
|  | met-d-Omega_6 | rs9616847 | 50868669 | T | A | 0.023821 | 0.388327 | 22 | 0.00415851 | 1.40E-08 | 114999 | NA |
|  | met-c-856 | rs11591147 | 55505647 | T | G | -0.309146 | 0.028839 | 1 | 0.039917 | 1.12E-14 | 13502 | 59.9807 |
|  | met-c-856 | rs190934192 | 55334001 | A | G | -0.243968 | 0.027571 | 1 | 0.043382 | 2.03E-08 | 13503 | 31.6262 |
|  | met-c-856 | rs7534572 | 62999675 | G | C | 0.083271 | 0.730699 | 1 | 0.01381 | 1.81E-09 | 13503 | 36.3580 |
|  | met-c-856 | rs1260326 | 27730940 | C | T | -0.07798 | 0.63681 | 2 | 0.012722 | 9.73E-10 | 13506 | 37.5713 |
|  | met-c-856 | rs76246956 | 74783906 | A | G | 0.225705 | 0.030794 | 4 | 0.040317 | 2.35E-08 | 13502 | 31.3405 |
|  | met-c-856 | rs144064722 | 73406173 | G | A | 0.23669 | 0.026411 | 4 | 0.039503 | 2.29E-09 | 13500 | 35.9004 |
|  | met-c-856 | rs79225634 | 74619639 | T | C | 0.08458 | 0.351533 | 5 | 0.012959 | 7.52E-11 | 13503 | 42.5983 |
|  | met-c-856 | rs3741298 | 116657561 | T | C | -0.14319 | 0.769753 | 11 | 0.01451 | 7.32E-23 | 13503 | 97.3847 |
|  | met-c-856 | rs174418 | 58687603 | C | T | -0.098225 | 0.562433 | 15 | 0.012554 | 5.99E-15 | 13504 | 61.2181 |
|  | met-c-856 | rs1800588 | 58723675 | T | C | 0.142532 | 0.249057 | 15 | 0.014482 | 9.47E-23 | 13504 | 96.8652 |
|  | met-c-856 | rs821840 | 56993886 | G | A | 0.08609 | 0.252132 | 16 | 0.014666 | 4.77E-09 | 13503 | 34.4574 |
|  | met-c-856 | rs10402112 | 11191677 | A | T | -0.166416 | 0.099716 | 19 | 0.021114 | 3.80E-15 | 13501 | 62.1225 |
|  | met-c-856 | rs7412 | 45412079 | T | C | -0.272206 | 0.057368 | 19 | 0.028286 | 8.06E-22 | 13501 | 92.6088 |
| Omega-6% | ebi-a-GCST90092935 | rs1168030 | 62968491 | T | C | -0.0492915 | 0.644929 | 1 | 0.00413738 | 1.00E-32 | 115006 | 141.9362 |
|  | ebi-a-GCST90092935 | rs6658257 | 93820684 | G | A | -0.0228432 | 0.602158 | 1 | 0.00405119 | 1.70E-08 | 115006 | 31.7943 |
|  | ebi-a-GCST90092935 | rs59484402 | 230302838 | CT | C | 0.027361 | 0.611698 | 1 | 0.00411002 | 2.80E-11 | 115006 | 44.3176 |
|  | ebi-a-GCST90092935 | rs182050989 | 27262545 | T | C | -0.0720426 | 0.02815 | 1 | 0.0119904 | 1.90E-09 | 115006 | 36.1004 |
|  | ebi-a-GCST90092935 | rs1260326 | 27730940 | C | T | 0.109743 | 0.604008 | 2 | 0.00404504 | 4.30E-162 | 115006 | 736.0512 |
|  | ebi-a-GCST90092935 | rs4665710 | 21221035 | C | A | -0.047267 | 0.792573 | 2 | 0.00486875 | 2.80E-22 | 115006 | 94.2499 |
|  | ebi-a-GCST90092935 | rs2972140 | 227104111 | C | T | -0.0340375 | 0.650628 | 2 | 0.00414245 | 2.10E-16 | 115006 | 67.5151 |
|  | ebi-a-GCST90092935 | rs13389219 | 165528876 | T | C | 0.038325 | 0.392574 | 2 | 0.00405363 | 3.20E-21 | 115006 | 89.3874 |
|  | ebi-a-GCST90092935 | rs78058190 | 219699999 | A | G | -0.0667368 | 0.050111 | 2 | 0.010245 | 7.30E-11 | 115006 | 42.4333 |
|  | ebi-a-GCST90092935 | rs684773 | 135956305 | C | A | -0.0347116 | 0.767069 | 3 | 0.00468622 | 1.30E-13 | 115006 | 54.8661 |
|  | ebi-a-GCST90092935 | rs1471251 | 87976359 | T | A | -0.0264073 | 0.397122 | 4 | 0.00406317 | 8.10E-11 | 115006 | 42.2394 |
|  | ebi-a-GCST90092935 | rs1316753 | 78531337 | C | G | 0.0228093 | 0.394862 | 5 | 0.00404888 | 1.80E-08 | 115006 | 31.7361 |
|  | ebi-a-GCST90092935 | rs11429307 | 55857025 | GT | G | -0.0445358 | 0.191495 | 5 | 0.00504741 | 1.10E-18 | 115006 | 77.8541 |
|  | ebi-a-GCST90092935 | rs117733303 | 160922870 | G | A | 0.209889 | 0.018512 | 6 | 0.014679 | 2.20E-46 | 115006 | 204.4497 |
|  | ebi-a-GCST90092935 | rs12212146 | 161125454 | C | T | -0.0567627 | 0.07432 | 6 | 0.00774962 | 2.40E-13 | 115006 | 53.6495 |
|  | ebi-a-GCST90092935 | rs28752523 | 32584772 | T | C | -0.0353026 | 0.193826 | 6 | 0.00500867 | 1.80E-12 | 115006 | 49.6785 |
|  | ebi-a-GCST90092935 | rs6938550 | 20462138 | A | G | 0.0399771 | 0.914146 | 6 | 0.00706603 | 1.50E-08 | 115006 | 32.0090 |
|  | ebi-a-GCST90092935 | rs10455872 | 161010118 | G | A | 0.138305 | 0.078987 | 6 | 0.0073371 | 2.90E-79 | 115006 | 355.3259 |
|  | ebi-a-GCST90092935 | rs6905288 | 43758873 | A | G | -0.02552 | 0.568514 | 6 | 0.00398662 | 1.50E-10 | 115006 | 40.9781 |
|  | ebi-a-GCST90092935 | rs199607859 | 139835418 | T | G | 0.0317367 | 0.594528 | 6 | 0.00404516 | 4.30E-15 | 115006 | 61.5534 |
|  | ebi-a-GCST90092935 | rs56001710 | 25983400 | T | A | 0.0335314 | 0.58059 | 7 | 0.00413696 | 5.30E-16 | 115006 | 65.6963 |
|  | ebi-a-GCST90092935 | rs3812316 | 73020337 | G | C | 0.0993677 | 0.129276 | 7 | 0.0058968 | 1.00E-63 | 115006 | 283.9603 |
|  | ebi-a-GCST90092935 | rs72555385 | 73123473 | G | A | -0.0540068 | 0.049314 | 7 | 0.0091908 | 4.20E-09 | 115006 | 34.5295 |
|  | ebi-a-GCST90092935 | rs11976955 | 130433594 | G | C | -0.0256733 | 0.314171 | 7 | 0.00427463 | 1.90E-09 | 115006 | 36.0717 |
|  | ebi-a-GCST90092935 | rs1561748 | 19733244 | C | G | 0.0494364 | 0.267445 | 8 | 0.00482644 | 1.30E-24 | 115006 | 104.9155 |
|  | ebi-a-GCST90092935 | rs328 | 19819724 | G | C | 0.137521 | 0.100282 | 8 | 0.00657047 | 2.80E-97 | 115006 | 438.0717 |
|  | ebi-a-GCST90092935 | rs4646246 | 18248661 | G | A | -0.028569 | 0.182591 | 8 | 0.00511872 | 2.40E-08 | 115006 | 31.1507 |
|  | ebi-a-GCST90092935 | rs28601761 | 126500031 | G | C | 0.0853476 | 0.420103 | 8 | 0.00405652 | 2.80E-98 | 115006 | 442.6652 |
|  | ebi-a-GCST90092935 | rs3860846 | 126631687 | T | C | -0.030052 | 0.275985 | 8 | 0.00447696 | 1.90E-11 | 115006 | 45.0589 |
|  | ebi-a-GCST90092935 | rs1736070 | 11665805 | T | C | 0.0259828 | 0.666408 | 8 | 0.00425289 | 1.00E-09 | 115006 | 37.3253 |
|  | ebi-a-GCST90092935 | rs295268 | 86429305 | C | T | 0.0280531 | 0.256162 | 9 | 0.00454438 | 6.70E-10 | 115006 | 38.1077 |
|  | ebi-a-GCST90092935 | rs10733306 | 16048248 | T | C | 0.0220496 | 0.460028 | 9 | 0.00400412 | 3.70E-08 | 115006 | 30.3241 |
|  | ebi-a-GCST90092935 | rs2068888 | 94839642 | A | G | 0.0257216 | 0.450146 | 10 | 0.0039792 | 1.00E-10 | 115006 | 41.7835 |
|  | ebi-a-GCST90092935 | rs12419462 | 27623309 | A | G | -0.0285755 | 0.228758 | 11 | 0.00471709 | 1.40E-09 | 115006 | 36.6977 |
|  | ebi-a-GCST90092935 | rs174528 | 61543499 | C | T | 0.0251335 | 0.377233 | 11 | 0.00408383 | 7.50E-10 | 115006 | 37.8766 |
|  | ebi-a-GCST90092935 | rs144018203 | 116916060 | C | G | -0.191837 | 0.010652 | 11 | 0.0200496 | 1.10E-21 | 115006 | 91.5489 |
|  | ebi-a-GCST90092935 | rs35169799 | 64031241 | T | C | -0.0485534 | 0.063323 | 11 | 0.00814674 | 2.50E-09 | 115006 | 35.5199 |
|  | ebi-a-GCST90092935 | rs964184 | 116648917 | C | G | 0.136188 | 0.867233 | 11 | 0.00583695 | 2.10E-120 | 115006 | 544.3845 |
|  | ebi-a-GCST90092935 | rs1047964 | 117156893 | T | G | 0.17031 | 0.010066 | 11 | 0.0201508 | 2.90E-17 | 115006 | 71.4325 |
|  | ebi-a-GCST90092935 | rs1546224 | 63696548 | T | C | -0.0236674 | 0.308641 | 12 | 0.00429622 | 3.60E-08 | 115006 | 30.3479 |
|  | ebi-a-GCST90092935 | rs73412716 | 112545197 | T | G | 0.0526186 | 0.069268 | 12 | 0.0079499 | 3.60E-11 | 115006 | 43.8082 |
|  | ebi-a-GCST90092935 | rs10773049 | 124506631 | C | T | 0.022457 | 0.393124 | 12 | 0.00405767 | 3.10E-08 | 115006 | 30.6302 |
|  | ebi-a-GCST90092935 | rs473224 | 58737341 | G | T | 0.0600834 | 0.853934 | 15 | 0.00567866 | 3.70E-26 | 115006 | 111.9482 |
|  | ebi-a-GCST90092935 | rs139974673 | 44027885 | C | T | -0.138506 | 0.025916 | 15 | 0.0124863 | 1.40E-28 | 115006 | 123.0466 |
|  | ebi-a-GCST90092935 | rs7402939 | 99183876 | C | T | -0.023501 | 0.623528 | 15 | 0.00413068 | 1.30E-08 | 115006 | 32.3690 |
|  | ebi-a-GCST90092935 | rs261290 | 58678720 | C | T | 0.0617512 | 0.654648 | 15 | 0.00417436 | 1.60E-49 | 115006 | 218.8321 |
|  | ebi-a-GCST90092935 | rs11508026 | 56999328 | T | C | 0.027324 | 0.432471 | 16 | 0.00401029 | 9.50E-12 | 115006 | 46.4234 |
|  | ebi-a-GCST90092935 | rs111351217 | 41163974 | A | G | -0.0497368 | 0.052245 | 17 | 0.00903706 | 3.70E-08 | 115006 | 30.2901 |
|  | ebi-a-GCST90092935 | rs72836561 | 41926126 | T | C | -0.0837853 | 0.031481 | 17 | 0.011342 | 1.50E-13 | 115006 | 54.5703 |
|  | ebi-a-GCST90092935 | rs1801689 | 64210580 | C | A | 0.0641739 | 0.03052 | 17 | 0.0115152 | 2.50E-08 | 115006 | 31.0580 |
|  | ebi-a-GCST90092935 | rs9908820 | 73408819 | G | A | -0.0279066 | 0.729127 | 17 | 0.00446315 | 4.00E-10 | 115006 | 39.0959 |
|  | ebi-a-GCST90092935 | rs58489806 | 19456917 | T | C | 0.070387 | 0.086416 | 19 | 0.00707508 | 2.60E-23 | 115006 | 98.9742 |
|  | ebi-a-GCST90092935 | rs56290633 | 45401783 | C | T | -0.0299572 | 0.53952 | 19 | 0.00428295 | 2.70E-12 | 115006 | 48.9234 |
|  | ebi-a-GCST90092935 | rs499765 | 49266390 | G | C | 0.0255461 | 0.338912 | 19 | 0.00422799 | 1.50E-09 | 115006 | 36.5074 |
|  | ebi-a-GCST90092935 | rs116843064 | 8429323 | A | G | 0.183791 | 0.01986 | 19 | 0.0142031 | 2.70E-38 | 115006 | 167.4489 |
|  | ebi-a-GCST90092935 | rs1065853 | 45413233 | T | G | -0.149197 | 0.080578 | 19 | 0.00730619 | 1.10E-92 | 115006 | 417.0021 |
|  | ebi-a-GCST90092935 | rs6073958 | 44551855 | C | T | -0.0474619 | 0.198745 | 20 | 0.00497167 | 1.30E-21 | 115006 | 91.1351 |
|  | met-d-Omega_6_pct | rs1168032 | 62967747 | G | A | -0.0496652 | 0.644939 | 1 | 0.00419905 | 1.40E-32 | 114999 | NA |
|  | met-d-Omega_6_pct | rs76890070 | 172190494 | T | C | 0.0674408 | 0.030697 | 1 | 0.0117769 | 4.30E-09 | 114999 | NA |
|  | met-d-Omega_6_pct | rs182050989 | 27262545 | T | C | -0.0727056 | 0.028151 | 1 | 0.0121688 | 8.30E-09 | 114999 | NA |
|  | met-d-Omega_6_pct | rs59484402 | 230302838 | C | CT | -0.0268674 | 0.388291 | 1 | 0.0041713 | 1.20E-10 | 114999 | NA |
|  | met-d-Omega_6_pct | rs1260326 | 27730940 | C | T | 0.109259 | 0.60401 | 2 | 0.00410526 | 7.00E-159 | 114999 | NA |
|  | met-d-Omega_6_pct | rs4665710 | 21221035 | C | A | -0.0470122 | 0.792578 | 2 | 0.00494139 | 3.70E-22 | 114999 | NA |
|  | met-d-Omega_6_pct | rs78058190 | 219699999 | A | G | -0.0677909 | 0.050105 | 2 | 0.0103987 | 2.20E-11 | 114999 | NA |
|  | met-d-Omega_6_pct | rs13389219 | 165528876 | T | C | 0.0383574 | 0.392567 | 2 | 0.00411407 | 2.90E-21 | 114999 | NA |
|  | met-d-Omega_6_pct | rs2943656 | 227121918 | G | A | -0.033583 | 0.635894 | 2 | 0.00416924 | 1.60E-15 | 114999 | NA |
|  | met-d-Omega_6_pct | rs534944 | 135961031 | G | T | -0.0344031 | 0.767256 | 3 | 0.00475995 | 3.90E-14 | 114999 | NA |
|  | met-d-Omega_6_pct | rs10020067 | 88181491 | A | C | -0.0268089 | 0.381451 | 4 | 0.00413478 | 5.60E-11 | 114999 | NA |
|  | met-d-Omega_6_pct | rs11429307 | 55857025 | G | GT | 0.0446837 | 0.808497 | 5 | 0.00512231 | 2.50E-18 | 114999 | NA |
|  | met-d-Omega_6_pct | rs199607859 | 139835418 | T | G | 0.0329128 | 0.594534 | 6 | 0.00410536 | 1.90E-15 | 114999 | NA |
|  | met-d-Omega_6_pct | rs6938550 | 20462138 | A | G | 0.0391913 | 0.914145 | 6 | 0.00717133 | 1.90E-08 | 114999 | NA |
|  | met-d-Omega_6_pct | rs28752523 | 32584772 | T | C | -0.0361874 | 0.193821 | 6 | 0.00508331 | 4.30E-13 | 114999 | NA |
|  | met-d-Omega_6_pct | rs6905288 | 43758873 | A | G | -0.0255665 | 0.56854 | 6 | 0.00404617 | 1.30E-10 | 114999 | NA |
|  | met-d-Omega_6_pct | rs117733303 | 160922870 | G | A | 0.215888 | 0.018513 | 6 | 0.0148972 | 2.20E-47 | 114999 | NA |
|  | met-d-Omega_6_pct | rs10455872 | 161010118 | G | A | 0.139161 | 0.078988 | 6 | 0.00744645 | 9.30E-79 | 114999 | NA |
|  | met-d-Omega_6_pct | rs12212146 | 161125454 | C | T | -0.0568652 | 0.074315 | 6 | 0.00786527 | 8.80E-14 | 114999 | NA |
|  | met-d-Omega_6_pct | rs72555385 | 73123473 | G | A | -0.0532093 | 0.049322 | 7 | 0.00932668 | 1.50E-08 | 114999 | NA |
|  | met-d-Omega_6_pct | rs11976955 | 130433594 | G | C | -0.0248228 | 0.314177 | 7 | 0.00433806 | 4.20E-09 | 114999 | NA |
|  | met-d-Omega_6_pct | rs56001710 | 25983400 | T | A | 0.0328314 | 0.580583 | 7 | 0.00419825 | 1.30E-15 | 114999 | NA |
|  | met-d-Omega_6_pct | rs3812316 | 73020337 | G | C | 0.0999463 | 0.129249 | 7 | 0.00598515 | 1.00E-62 | 114999 | NA |
|  | met-d-Omega_6_pct | rs4646246 | 18248661 | G | A | -0.0283178 | 0.18259 | 8 | 0.00519496 | 8.80E-09 | 114999 | NA |
|  | met-d-Omega_6_pct | rs1561748 | 19733244 | C | G | 0.0483024 | 0.267454 | 8 | 0.0048983 | 6.40E-22 | 114999 | NA |
|  | met-d-Omega_6_pct | rs28601761 | 126500031 | G | C | 0.0851115 | 0.420119 | 8 | 0.00411688 | 5.70E-97 | 114999 | NA |
|  | met-d-Omega_6_pct | rs1736070 | 11665805 | T | C | 0.0245965 | 0.666409 | 8 | 0.0043162 | 2.00E-08 | 114999 | NA |
|  | met-d-Omega_6_pct | rs4871624 | 126629328 | G | T | -0.0289405 | 0.287462 | 8 | 0.00446476 | 1.40E-10 | 114999 | NA |
|  | met-d-Omega_6_pct | rs328 | 19819724 | G | C | 0.135169 | 0.100283 | 8 | 0.00666822 | 4.40E-93 | 114999 | NA |
|  | met-d-Omega_6_pct | rs7047907 | 86368660 | A | G | -0.0259918 | 0.742524 | 9 | 0.00461138 | 1.30E-08 | 114999 | NA |
|  | met-d-Omega_6_pct | rs10733306 | 16048248 | T | C | 0.0227079 | 0.46003 | 9 | 0.0040634 | 3.70E-08 | 114999 | NA |
|  | met-d-Omega_6_pct | rs2068888 | 94839642 | A | G | 0.0255934 | 0.450134 | 10 | 0.0040382 | 2.80E-10 | 114999 | NA |
|  | met-d-Omega_6_pct | rs174528 | 61543499 | C | T | 0.0248308 | 0.377235 | 11 | 0.00414455 | 1.10E-09 | 114999 | NA |
|  | met-d-Omega_6_pct | rs141078087 | 27631005 | A | AT | 0.0288468 | 0.772246 | 11 | 0.00481299 | 2.20E-10 | 114999 | NA |
|  | met-d-Omega_6_pct | rs35169799 | 64031241 | T | C | -0.0478563 | 0.063327 | 11 | 0.00826765 | 1.70E-09 | 114999 | NA |
|  | met-d-Omega_6_pct | rs964184 | 116648917 | C | G | 0.13521 | 0.867229 | 11 | 0.00592342 | 7.29E-117 | 114999 | NA |
|  | met-d-Omega_6_pct | rs144018203 | 116916060 | C | G | -0.189872 | 0.010653 | 11 | 0.0203471 | 3.80E-22 | 114999 | NA |
|  | met-d-Omega_6_pct | rs1047964 | 117156893 | T | G | 0.171083 | 0.010067 | 11 | 0.0204498 | 9.50E-17 | 114999 | NA |
|  | met-d-Omega_6_pct | rs1546224 | 63696548 | T | C | -0.0239486 | 0.308646 | 12 | 0.00435994 | 4.80E-08 | 114999 | NA |
|  | met-d-Omega_6_pct | rs1716407 | 124515218 | A | G | -0.0230595 | 0.592755 | 12 | 0.00409177 | 1.70E-08 | 114999 | NA |
|  | met-d-Omega_6_pct | rs73412716 | 112545197 | T | G | 0.0539254 | 0.069272 | 12 | 0.00806764 | 3.50E-11 | 114999 | NA |
|  | met-d-Omega_6_pct | rs473224 | 58737341 | G | T | 0.0606142 | 0.853925 | 15 | 0.00576268 | 1.10E-25 | 114999 | NA |
|  | met-d-Omega_6_pct | rs139974673 | 44027885 | C | T | -0.137656 | 0.025918 | 15 | 0.012671 | 9.10E-28 | 114999 | NA |
|  | met-d-Omega_6_pct | rs261290 | 58678720 | C | T | 0.0629069 | 0.654653 | 15 | 0.00423625 | 2.10E-50 | 114999 | NA |
|  | met-d-Omega_6_pct | rs708272 | 56996288 | A | G | 0.0261942 | 0.432765 | 16 | 0.00405573 | 3.10E-11 | 114999 | NA |
|  | met-d-Omega_6_pct | rs72836561 | 41926126 | T | C | -0.083998 | 0.031483 | 17 | 0.0115096 | 5.80E-14 | 114999 | NA |
|  | met-d-Omega_6_pct | rs9908820 | 73408819 | G | A | -0.0281089 | 0.729141 | 17 | 0.00452935 | 1.70E-09 | 114999 | NA |
|  | met-d-Omega_6_pct | rs111351217 | 41163974 | A | G | -0.0534972 | 0.052244 | 17 | 0.00917078 | 1.60E-08 | 114999 | NA |
|  | met-d-Omega_6_pct | rs1801689 | 64210580 | C | A | 0.0649827 | 0.030526 | 17 | 0.0116846 | 6.90E-09 | 114999 | NA |
|  | met-d-Omega_6_pct | rs56290633 | 45401783 | C | T | -0.029505 | 0.539528 | 19 | 0.00434643 | 3.10E-11 | 114999 | NA |
|  | met-d-Omega_6_pct | rs499765 | 49266390 | G | C | 0.0262628 | 0.338914 | 19 | 0.00429063 | 1.70E-09 | 114999 | NA |
|  | met-d-Omega_6_pct | rs8107974 | 19388500 | T | A | 0.0726822 | 0.075537 | 19 | 0.00761905 | 1.80E-21 | 114999 | NA |
|  | met-d-Omega_6_pct | rs1065853 | 45413233 | T | G | -0.148377 | 0.080578 | 19 | 0.00741446 | 1.60E-89 | 114999 | NA |
|  | met-d-Omega_6_pct | rs116843064 | 8429323 | A | G | 0.183686 | 0.019861 | 19 | 0.0144132 | 4.50E-37 | 114999 | NA |
|  | met-d-Omega_6_pct | rs4810479 | 44545048 | T | C | 0.0440519 | 0.75018 | 20 | 0.00464348 | 2.60E-22 | 114999 | NA |
|  | met-d-Omega_6_pct | rs2835056 | 37005665 | G | A | 0.0217796 | 0.345823 | 21 | 0.00421981 | 3.90E-08 | 114999 | NA |
| Omega-6：Omega-3 | ebi-a-GCST90092934 | rs6693447 | 2330190 | G | T | -0.0250996 | 0.461701 | 1 | 0.00398654 | 3.10E-10 | 115006 | 39.6407 |
|  | ebi-a-GCST90092934 | rs583609 | 62916796 | C | T | 0.0428152 | 0.352508 | 1 | 0.0041587 | 7.40E-25 | 115006 | 105.9939 |
|  | ebi-a-GCST90092934 | rs1260326 | 27730940 | C | T | 0.066446 | 0.604008 | 2 | 0.00405986 | 3.30E-60 | 115006 | 267.8648 |
|  | ebi-a-GCST90092934 | rs4860987 | 69491284 | T | A | -0.0411323 | 0.258612 | 4 | 0.00479927 | 1.00E-17 | 115006 | 73.4540 |
|  | ebi-a-GCST90092934 | rs10075801 | 131677642 | G | A | -0.02358 | 0.485487 | 5 | 0.00401401 | 4.20E-09 | 115006 | 34.5089 |
|  | ebi-a-GCST90092934 | rs2394976 | 31311912 | T | G | 0.0315075 | 0.161642 | 6 | 0.00538488 | 4.90E-09 | 115006 | 34.2354 |
|  | ebi-a-GCST90092934 | rs62466318 | 73042085 | T | C | 0.0599701 | 0.204201 | 7 | 0.00494834 | 8.40E-34 | 115006 | 146.8759 |
|  | ebi-a-GCST90092934 | rs4000713 | 25990597 | A | G | 0.0302187 | 0.295416 | 7 | 0.00435921 | 4.10E-12 | 115006 | 48.0547 |
|  | ebi-a-GCST90092934 | rs112875651 | 126506694 | A | G | 0.0719492 | 0.392337 | 8 | 0.00412529 | 4.00E-68 | 115006 | 304.1886 |
|  | ebi-a-GCST90092934 | rs7819706 | 19844415 | G | A | 0.0363774 | 0.118288 | 8 | 0.00614755 | 3.30E-09 | 115006 | 35.0154 |
|  | ebi-a-GCST90092934 | rs7916868 | 64988931 | T | A | -0.0282856 | 0.504504 | 10 | 0.00397492 | 1.10E-12 | 115006 | 50.6377 |
|  | ebi-a-GCST90092934 | rs9332238 | 96748492 | A | G | -0.0347177 | 0.198299 | 10 | 0.00498195 | 3.20E-12 | 115006 | 48.5627 |
|  | ebi-a-GCST90092934 | rs150370599 | 60718792 | T | C | -0.0475725 | 0.080318 | 11 | 0.00728355 | 6.50E-11 | 115006 | 42.6605 |
|  | ebi-a-GCST90092934 | rs174564 | 61588305 | G | A | 0.371159 | 0.347009 | 11 | 0.00414236 | 1.00E-200 | 115006 | 8028.3133 |
|  | ebi-a-GCST90092934 | rs77323894 | 60896505 | C | G | -0.0682803 | 0.038434 | 11 | 0.0107365 | 2.00E-10 | 115006 | 40.4451 |
|  | ebi-a-GCST90092934 | rs12226389 | 61823630 | C | T | 0.0592832 | 0.185805 | 11 | 0.00512548 | 6.10E-31 | 115006 | 133.7809 |
|  | ebi-a-GCST90092934 | rs673335 | 75450576 | C | T | 0.0611093 | 0.159752 | 11 | 0.00541161 | 1.40E-29 | 115006 | 127.5151 |
|  | ebi-a-GCST90092934 | rs11230829 | 61701898 | G | A | 0.103011 | 0.027785 | 11 | 0.0144741 | 1.10E-12 | 115006 | 50.6505 |
|  | ebi-a-GCST90092934 | rs145786300 | 61406089 | A | G | 0.176634 | 0.011993 | 11 | 0.0187777 | 5.10E-21 | 115006 | 88.4838 |
|  | ebi-a-GCST90092934 | rs195445 | 61744342 | T | C | 0.0344283 | 0.688414 | 11 | 0.00427728 | 8.30E-16 | 115006 | 64.7882 |
|  | ebi-a-GCST90092934 | rs964184 | 116648917 | C | G | 0.0746316 | 0.867233 | 11 | 0.00582544 | 1.40E-37 | 115006 | 164.1300 |
|  | ebi-a-GCST90092934 | rs139974673 | 44027885 | C | T | -0.117068 | 0.025916 | 15 | 0.0125202 | 8.70E-21 | 115006 | 87.4287 |
|  | ebi-a-GCST90092934 | rs1077835 | 58723426 | G | A | -0.0998071 | 0.219711 | 15 | 0.00480305 | 6.60E-96 | 115006 | 431.8060 |
|  | ebi-a-GCST90092934 | rs1532085 | 58683366 | G | A | 0.0862885 | 0.613968 | 15 | 0.00407981 | 2.80E-99 | 115006 | 447.3279 |
|  | ebi-a-GCST90092934 | rs72789541 | 15127534 | A | T | 0.0906088 | 0.295973 | 16 | 0.00435849 | 5.40E-96 | 115006 | 432.1840 |
|  | ebi-a-GCST90092934 | rs35390787 | 15678414 | CA | C | -0.0255265 | 0.438254 | 16 | 0.00409167 | 4.40E-10 | 115006 | 38.9208 |
|  | ebi-a-GCST90092934 | rs8074191 | 17407191 | C | T | 0.0281472 | 0.756121 | 17 | 0.00463331 | 1.20E-09 | 115006 | 36.9052 |
|  | ebi-a-GCST90092934 | rs2463523 | 43671737 | C | A | 0.0401451 | 0.23479 | 17 | 0.00475753 | 3.20E-17 | 115006 | 71.2036 |
|  | ebi-a-GCST90092934 | rs9947684 | 47166694 | G | A | -0.0276606 | 0.654466 | 18 | 0.00417279 | 3.40E-11 | 115006 | 43.9410 |
|  | ebi-a-GCST90092934 | rs116843064 | 8429323 | A | G | 0.0882813 | 0.01986 | 19 | 0.014248 | 5.80E-10 | 115006 | 38.3910 |
|  | ebi-a-GCST90092934 | rs12976395 | 45441907 | C | G | -0.0272184 | 0.504524 | 19 | 0.0042714 | 1.90E-10 | 115006 | 40.6055 |
|  | ebi-a-GCST90092934 | rs1065853 | 45413233 | T | G | -0.0803838 | 0.080578 | 19 | 0.00732929 | 5.50E-28 | 115006 | 120.2855 |
|  | ebi-a-GCST90092934 | rs58542926 | 19379549 | T | C | 0.144499 | 0.074379 | 19 | 0.00757644 | 4.30E-81 | 115006 | 363.7469 |
|  | ebi-a-GCST90092934 | rs182611493 | 19458388 | G | A | 0.19692 | 0.012519 | 19 | 0.0191342 | 7.70E-25 | 115006 | 105.9154 |
|  | ebi-a-GCST90092934 | rs117143374 | 40555561 | C | T | 0.0318136 | 0.14225 | 21 | 0.00571328 | 2.60E-08 | 115006 | 31.0066 |
|  | met-d-Omega_6_by_Omega_3 | rs6698680 | 2329661 | G | A | -0.0258639 | 0.461958 | 1 | 0.00410413 | 2.10E-11 | 114999 | NA |
|  | met-d-Omega_6_by_Omega_3 | rs638714 | 62906489 | T | G | 0.0438817 | 0.345963 | 1 | 0.00431435 | 3.70E-25 | 114999 | NA |
|  | met-d-Omega_6_by_Omega_3 | rs1260326 | 27730940 | C | T | 0.0654544 | 0.60401 | 2 | 0.004179 | 4.20E-55 | 114999 | NA |
|  | met-d-Omega_6_by_Omega_3 | rs4860987 | 69491284 | T | A | -0.0426258 | 0.258609 | 4 | 0.00493905 | 6.80E-18 | 114999 | NA |
|  | met-d-Omega_6_by_Omega_3 | rs11242109 | 131677047 | T | G | -0.0239824 | 0.479016 | 5 | 0.00408984 | 1.40E-09 | 114999 | NA |
|  | met-d-Omega_6_by_Omega_3 | rs2394976 | 31311912 | T | G | 0.0339337 | 0.161648 | 6 | 0.00554242 | 5.80E-10 | 114999 | NA |
|  | met-d-Omega_6_by_Omega_3 | rs1564348 | 160578860 | C | T | 0.030467 | 0.169745 | 6 | 0.00544576 | 1.40E-08 | 114999 | NA |
|  | met-d-Omega_6_by_Omega_3 | rs4000713 | 25990597 | A | G | 0.0308836 | 0.295408 | 7 | 0.00448656 | 6.10E-13 | 114999 | NA |
|  | met-d-Omega_6_by_Omega_3 | rs62466318 | 73042085 | T | C | 0.0606132 | 0.204178 | 7 | 0.00509341 | 4.40E-33 | 114999 | NA |
|  | met-d-Omega_6_by_Omega_3 | rs73109460 | 44785800 | A | G | 0.0338895 | 0.123622 | 7 | 0.00625247 | 2.20E-09 | 114999 | NA |
|  | met-d-Omega_6_by_Omega_3 | rs112875651 | 126506694 | A | G | 0.0718479 | 0.392346 | 8 | 0.00424549 | 6.50E-65 | 114999 | NA |
|  | met-d-Omega_6_by_Omega_3 | rs13273454 | 19940058 | T | C | 0.0233472 | 0.470545 | 8 | 0.00411509 | 5.80E-09 | 114999 | NA |
|  | met-d-Omega_6_by_Omega_3 | rs10733306 | 16048248 | T | C | 0.0225443 | 0.46003 | 9 | 0.00413317 | 3.20E-08 | 114999 | NA |
|  | met-d-Omega_6_by_Omega_3 | rs7916868 | 64988931 | T | A | -0.0295041 | 0.504491 | 10 | 0.00409087 | 5.10E-14 | 114999 | NA |
|  | met-d-Omega_6_by_Omega_3 | rs55891451 | 96728169 | C | A | -0.0355372 | 0.201728 | 10 | 0.0051102 | 1.20E-12 | 114999 | NA |
|  | met-d-Omega_6_by_Omega_3 | rs174564 | 61588305 | G | A | 0.371404 | 0.347013 | 11 | 0.00426515 | 1.00E-200 | 114999 | NA |
|  | met-d-Omega_6_by_Omega_3 | rs964184 | 116648917 | C | G | 0.0735236 | 0.867229 | 11 | 0.00599779 | 5.30E-35 | 114999 | NA |
|  | met-d-Omega_6_by_Omega_3 | rs673335 | 75450576 | C | T | 0.0598626 | 0.159762 | 11 | 0.00557189 | 7.40E-28 | 114999 | NA |
|  | met-d-Omega_6_by_Omega_3 | rs2232143 | 60899701 | C | T | -0.10962 | 0.021879 | 11 | 0.0145796 | 9.40E-14 | 114999 | NA |
|  | met-d-Omega_6_by_Omega_3 | rs145659493 | 61850279 | A | C | -0.115733 | 0.015854 | 11 | 0.0163525 | 4.30E-13 | 114999 | NA |
|  | met-d-Omega_6_by_Omega_3 | rs143355652 | 61453822 | T | C | 0.16027 | 0.010467 | 11 | 0.0205193 | 7.60E-15 | 114999 | NA |
|  | met-d-Omega_6_by_Omega_3 | rs11230829 | 61701898 | G | A | 0.102081 | 0.027786 | 11 | 0.0149026 | 4.00E-12 | 114999 | NA |
|  | met-d-Omega_6_by_Omega_3 | rs12226389 | 61823630 | C | T | 0.0563519 | 0.18582 | 11 | 0.00527721 | 8.40E-27 | 114999 | NA |
|  | met-d-Omega_6_by_Omega_3 | rs145786300 | 61406089 | A | G | 0.174042 | 0.011985 | 11 | 0.0193409 | 1.10E-19 | 114999 | NA |
|  | met-d-Omega_6_by_Omega_3 | rs149820547 | 61983775 | G | T | 0.0653372 | 0.042103 | 11 | 0.0102102 | 1.20E-10 | 114999 | NA |
|  | met-d-Omega_6_by_Omega_3 | rs139974673 | 44027885 | C | T | -0.117385 | 0.025918 | 15 | 0.0128859 | 9.10E-21 | 114999 | NA |
|  | met-d-Omega_6_by_Omega_3 | rs1560390 | 58580781 | C | T | 0.0352816 | 0.219684 | 15 | 0.00496671 | 2.80E-13 | 114999 | NA |
|  | met-d-Omega_6_by_Omega_3 | rs261291 | 58680178 | C | T | -0.0891379 | 0.356068 | 15 | 0.00427499 | 9.90E-99 | 114999 | NA |
|  | met-d-Omega_6_by_Omega_3 | rs11632618 | 58724706 | A | G | -0.0815347 | 0.069927 | 15 | 0.00800529 | 1.20E-24 | 114999 | NA |
|  | met-d-Omega_6_by_Omega_3 | rs35390787 | 15678414 | C | CA | 0.0252031 | 0.561747 | 16 | 0.0042113 | 9.70E-10 | 114999 | NA |
|  | met-d-Omega_6_by_Omega_3 | rs72789541 | 15127534 | A | T | 0.0881716 | 0.29597 | 16 | 0.00448603 | 1.40E-86 | 114999 | NA |
|  | met-d-Omega_6_by_Omega_3 | rs16940904 | 44186063 | T | C | 0.0412048 | 0.226571 | 17 | 0.00490538 | 2.30E-18 | 114999 | NA |
|  | met-d-Omega_6_by_Omega_3 | rs7222755 | 73888423 | G | A | 0.0239677 | 0.290719 | 17 | 0.00450454 | 4.10E-08 | 114999 | NA |
|  | met-d-Omega_6_by_Omega_3 | rs8074191 | 17407191 | C | T | 0.0277573 | 0.756127 | 17 | 0.00476891 | 4.80E-09 | 114999 | NA |
|  | met-d-Omega_6_by_Omega_3 | rs9947684 | 47166694 | G | A | -0.0283663 | 0.654493 | 18 | 0.00429465 | 5.70E-12 | 114999 | NA |
|  | met-d-Omega_6_by_Omega_3 | rs1065853 | 45413233 | T | G | -0.0791361 | 0.080578 | 19 | 0.00754362 | 1.20E-26 | 114999 | NA |
|  | met-d-Omega_6_by_Omega_3 | rs12976395 | 45441907 | C | G | -0.0274747 | 0.504524 | 19 | 0.00439626 | 7.50E-10 | 114999 | NA |
|  | met-d-Omega_6_by_Omega_3 | rs116843064 | 8429323 | A | G | 0.0839559 | 0.019861 | 19 | 0.0146642 | 1.60E-08 | 114999 | NA |
|  | met-d-Omega_6_by_Omega_3 | rs58542926 | 19379549 | T | C | 0.143289 | 0.074383 | 19 | 0.00779781 | 3.40E-79 | 114999 | NA |
|  | met-d-Omega_6_by_Omega_3 | rs182611493 | 19458388 | G | A | 0.18661 | 0.012519 | 19 | 0.0196926 | 1.10E-21 | 114999 | NA |
|  | met-d-Omega_6_by_Omega_3 | rs117143374 | 40555561 | C | T | 0.0343687 | 0.142254 | 21 | 0.00587978 | 6.00E-09 | 114999 | NA |

Chr,chromosome; Eaf,Effect Allele Frequency; NA,Not Available; SNPs, Single-nucleotide polymorphisms; Se,standard error

Table S3 Overview of the used datasets.

| **Expoure or outcome** | **GWAS ID** | **Name** | **Data source** | **Year** | **Ncase** | **Ncontrol** | **Samplesize** | **Number of SNPs** |
| --- | --- | --- | --- | --- | --- | --- | --- | --- |
| Exposure | ebi-a-GCST90092931 | Omega-3 fatty acid levels | IEU | 2022 | NA | NA | 115,006 | 11,590,399 |
| Exposure | met-d-Omega_3 | Omega-3 fatty acids | IEU | 2020 | NA | NA | 114,999 | 12,321,875 |
| Exposure | met-c-855 | Omega-3 fatty acids | IEU | 2016 | NA | NA | 13,544 | 11,401,623 |
| Exposure | ebi-a-GCST90092932 | Ratio of omega-3 fatty acids to total fatty acids | IEU | 2022 | NA | NA | 115,006 | 11,590,399 |
| Exposure | met-d-Omega_3_pct | Ratio of omega-3 fatty acids to total fatty acids | IEU | 2020 | NA | NA | 114,999 | 12,321,875 |
| Exposure | ebi-a-GCST90092933 | Omega-6 fatty acid levels | IEU | 2022 | NA | NA | 115,006 | 11,590,399 |
| Exposure | met-d-Omega_6 | Omega-6 fatty acids | IEU | 2020 | NA | NA | 114,999 | 12,321,875 |
| Exposure | met-c-856 | Omega-6 fatty acids | IEU | 2016 | NA | NA | 13,506 | 11,398,837 |
| Exposure | ebi-a-GCST90092935 | Ratio of omega-6 fatty acids to total fatty acids | IEU | 2022 | NA | NA | 115,006 | 11,590,399 |
| Exposure | met-d-Omega_6_pct | Ratio of omega-6 fatty acids to total fatty acids | IEU | 2020 | NA | NA | 114,999 | 12,321,875 |
| Exposure | ebi-a-GCST90092934 | Ratio of omega-6 fatty acids to omega-3 fatty acids | IEU | 2022 | NA | NA | 115,006 | 11,590,399 |
| Exposure | met-d-Omega_6_by_Omega_3 | Ratio of omega-6 fatty acids to omega-3 fatty acids | IEU | 2020 | NA | NA | 114,999 | 12,321,875 |
| Outcome | G6_ALZHEIMER | Alzheimer disease | FINN | 2022 | 9301 | 367976 | NA | 15 |
| Outcome | AD | Alzheimer's disease | UKB | NA | NA | NA | NA | NA |
| Outcome | F5_ADHD | Disturbance of activity and attention | FINN | 2022 | 2340 | 371117 |  | 682 |
| Outcome | ebi-a-GCST005362 | Attention deficit hyperactivity disorder | IEU | 2017 | 14154 | 17948 | 32102 | 7414807 |
| Outcome | ebi-a-GCST012597 | Attention deficit hyperactivity disorder | IEU | 2017 | 4945 | 16246 | 21191 | 7392559 |
| Outcome | ieu-a-1183 | ADHD | IEU | 2017 | 20183 | 35191 | 55374 | 8047420 |
| Outcome | F5_SCHZPHR | Schizophrenia | FINN | 2022 | 6515 | 364160 | NA | 592 |
| Outcome | F20 | Diagnoses - main ICD10: F20 Schizophrenia | UKB | NA | NA | NA | NA | NA |
| Outcome | 20002_1289 | Non-cancer illness code, self-reported: schizophrenia | UKB | 2023 | NA | NA | 393104 | NA |
| Outcome | 20544_2 | Mental health problems ever diagnosed by a professional: Schizophrenia | UKB | 2017 | NA | NA | 50068 | NA |
| Outcome | ebi-a-GCST90018919 | Schizophrenia | IEU | 2021 | 6334 | 445120 | 451454 | 24192920 |
| Outcome | ebi-a-GCST90018699 | Schizophrenia | IEU | 2021 | 99 | 177794 | 177893 | 12454529 |
| Outcome | ieu-b-5099 | Schizophrenia | IEU | 2022 | 76755 | 243649 | 320404 | NA |
| Outcome | ieu-b-5100 | Schizophrenia | IEU | 2022 | 64322 | 90947 | 155269 | NA |
| Outcome | ieu-b-5102 | Schizophrenia | IEU | 2022 | 52017 | 75889 | 127906 | NA |
| Outcome | ieu-a-22 | Schizophrenia | IEU | 2014 | 35476 | 46839 | 82315 | 9444231 |
| Outcome | ieu-b-42 | schizophrenia | IEU | 2014 | 33640 | 43456 | 77096 | 15358497 |
| Outcome | ieu-b-5070 | Schizophrenia | IEU | 2019 | 22778 | 35362 | 58140 | 10663823 |
| Outcome | ieu-b-5101 | Schizophrenia | IEU | 2022 | 12305 | 15058 | 27363 | NA |
| Outcome | ieu-b-5098 | Schizophrenia | IEU | 2022 | 5998 | 3826 | 9824 | NA |
| Outcome | ieu-b-5103 | Schizophrenia | IEU | 2022 | 1234 | 3090 | 3090 | NA |
| Outcome | ieu-a-810 | Schizophrenia | IEU | NA | 954 | 1195 | 2149 | 809849 |
| Outcome | F5_PTSD | Post-traumatic stress disorder | FINN | 2022 | 2282 | 337577 | NA | 554 |
| Outcome | 20002_1469 | Non-cancer illness code, self-reported: post-traumatic stress disorder | UKB | 2023 | NA | NA | 393104 | NA |
| Outcome | F5_MANIA | Manic episode | FINN | 2022 | 934 | 329192 | NA | 640 |
| Outcome | F5_DEPRESSIO | Depression | FINN | 2022 | 43280 | 329192 | NA | 50 |
| Outcome | F5_DEPRESSIO | Depression | UKB | NA | NA | NA | NA | NA |
| Outcome | 20002_1286 | Non-cancer illness code, self-reported: depression | UKB | 2023 | NA | NA | 393104 | NA |
| Outcome | 20544_11 | Mental health problems ever diagnosed by a professional: Depression | UKB | 2017 | NA | NA | 50068 | NA |
| Outcome | 20544_11 | Mental health problems ever diagnosed by a professional: Depression | UKB | 2017 | NA | NA | 50068 | NA |
| Outcome | ebi-a-GCST009979 | Major depressive disorder | IEU | 2020 | 29475 | 63482 | 92957 | 7767934 |
| Outcome | ebi-a-GCST90086058 | Major depressive disorder | IEU | 2021 | 7264 | 49373 | 56637 | 14426915 |
| Outcome | ebi-a-GCST90086059 | Major depressive disorder | IEU | 2021 | 7264 | 49373 | 56637 | 11498420 |
| Outcome | ebi-a-GCST90086060 | Major depressive disorder | IEU | 2021 | 7264 | 49373 | 56637 | 14462316 |
| Outcome | ebi-a-GCST90086061 | Major depressive disorder | IEU | 2021 | 7264 | 49373 | 56637 | 10810543 |
| Outcome | ebi-a-GCST90086062 | Major depressive disorder | IEU | 2021 | 7264 | 49373 | 56637 | 14462316 |
| Outcome | ieu-a-804 | Major depressive disorder | IEU | 2013 | 9240 | 9519 | 18759 | 123041 |
| Outcome | ieu-a-805 | Major depressive disorder | IEU | 2013 | 9240 | 9519 | 18759 | 1235110 |
| Outcome | ieu-a-1188 | Major Depressive Disorder | IEU | 2018 | 59851 | 113154 | 173005 | 13554550 |
| Outcome | F5_BIPO | Bipolar affective disorders | FINN | 2022 | 7006 | 329192 | NA | 582 |
| Outcome | F31 | Diagnoses - main ICD10: F31 Bipolar affective disorder | UKB | NA | NA | NA | NA | NA |
| Outcome | KRA_PSY_AnxietyIETY_EXMORE | Anxietyiety disorders (more control exclusions) | FINN | 2022 | 40191 | 277526 | NA | 120 |
| Outcome | F5_ALLAnxietyIOUS | All Anxietyiety disorders | FINN | 2022 | 24662 | 337577 | NA | 194 |
| Outcome | F5_ALLAnxietyIOUS | All Anxietyiety disorders | UKB | NA | NA | NA | NA | NA |
| Outcome | KRA_PSY_AnxietyIETY | Anxietyiety disorders | UKB | NA | NA | NA | NA | NA |
| Outcome | F5_OCD | Obsessive-compulsive disorder | FINN | 2022 | 1962 | 337577 | NA | 586 |
| Outcome | 20544_7 | Mental health problems ever diagnosed by a professional: Obsessive compulsive disorder (OCD) | UKB | 2017 | NA | NA | 50068 | NA |
| Outcome | 20544_7 | Mental health problems ever diagnosed by a professional: Obsessive compulsive disorder (OCD) | UKB | 2017 | NA | NA | 50068 | NA |
| Outcome | 20544_14 | Mental health problems ever diagnosed by a professional: Autism, Asperger's or autistic spectrum disorder | UKB | 2017 | NA | NA | 50068 | NA |
| Outcome | 20544_14 | Mental health problems ever diagnosed by a professional: Autism, Asperger's or autistic spectrum disorder | UKB | 2017 | NA | NA | 50068 | NA |
| Outcome | ieu-a-1185 | Autism Spectrum Disorder | IEU | 2017 | 18382 | 27969 | 46351 | 9112386 |
| Outcome | ieu-a-1184 | Autism Spectrum Disorder | IEU | 2015 | 5305 | 5305 | 10610 | 9499589 |
| Outcome | KRA_PSY_PERSON_EXMORE | Personality disorders (more control exclusions) | FINN | 2022 | 10012 | 277522 | NA | 339 |
| Outcome | KRA_PSY_PERSON | Personality disorders | UKB | NA | NA | NA | NA | NA |
| Outcome | 20544_4 | Mental health problems ever diagnosed by a professional: A personality disorder | UKB | 2017 | NA | NA | 50068 | NA |
| Outcome | 20544_4 | Mental health problems ever diagnosed by a professional: A personality disorder | UKB | 2017 | NA | NA | 50068 | NA |
| Outcome | F5_ANAPER | Anankastic personality disorder | FINN | 2022 | 1032 | 366637 | NA | 625 |
| Outcome | F5_AnxietyPER | Anxietyious personality disorder | FINN | 2022 | 534 | 366637 | NA | 580 |
| Outcome | F5_DEPPER | Dependent personality disorder | FINN | 2022 | 610 | 366637 | NA | 719 |
| Outcome | F5_DISPER | Dissocial personality disorder | FINN | 2022 | 467 | 366637 | NA | 792 |
| Outcome | F5_EMOPER | Emotionally unstable personality disorder | FINN | 2022 | 4183 | 366637 | NA | 508 |
| Outcome | F5_HISPER | Histrionic personality disorder | FINN | 2022 | 142 | 366637 | NA | 834 |
| Outcome | F5_MIXPER | Mixed and other personality disorders | FINN | 2022 | 2619 | 366637 | NA | 601 |
| Outcome | F5_PARAPER | Paranoid personality disorder | FINN | 2022 | 559 | 366637 | NA | 691 |
| Outcome | F5_SCHIZPER | Schizoid personality disorder | FINN | 2022 | 550 | 366637 | NA | 709 |

NA,Not available; SNP,single-nucleotide polymorphism

Table S4 Baseline characteristics of included studies in the meta-analysis.

| **References (author,year)** | **Country** | **Exposure** | **Data source** | **Ncontrol** | **Number of SNPs** | **Outcome** | **Data source** | **N case** | **N control** | **Samplesize** |
| --- | --- | --- | --- | --- | --- | --- | --- | --- | --- | --- |
| Zhang et al., (2023) | China | Omega-3、Omega-6、Omega-6：Omega-3 | UKB | 114 999 | 12 321 875 | BD | EBI | NR | NR | 34 950 |
| Xu et al., (2023) | USA | Omega-3%、Omega-6%、Omega-6：Omega-3 | IEU | 114999 | 10568861 | ADHD | PGC | 19099 | 34194 | 53293 |
|  |  |  |  |  |  | Anxiety | PGC | 7016 | 14745 | 21761 |
|  |  |  |  |  |  | ASD | PGC | 7016 | 14745 | 21761 |
|  |  |  |  |  |  | BD | PGC | 41917 | 371549 | 413466 |
|  |  |  |  |  |  | MDD | PGC | 170756 | 329443 | 500199 |
|  |  |  |  |  |  | OCD | PGC | 2688 | 7037 | 9725 |
|  |  |  |  |  |  | PTSD | PGC | 23212 | 151447 | 174659 |
|  |  |  |  |  |  | SCZ | PGC | 35476 | 46839 | 82315 |
| Milaneschi et al., (2019) | Netherlands | Omega-3 | MAGNETIC NMR GWAS | 24 925 | 6 | MDD | PGC | 135 458 | 344901 | NR |
| Ma et al., (2023) | China | Omega-3 | Kettunen et al.,2016 | 13544 | 11 401 622 | MDD | IEU | 59 851 | 113 154 | 17 3005 |
| Zeng et al., (2022) | China | Omega-3 | UKB | 114999 | 49 | Depression | PGC | 246363 | 561190 | NR |
|  |  | Omega-6 |  |  | 64 |  |  |  |  |  |
| Rebecca et al., (2022) | UK | Omega-3 | UKB | 115078 | 43 | MDD | PGC | 135458 | 344901 | 480359 |
|  |  | Omega-3% |  |  | 33 |  |  |  |  |  |
|  |  | Omega-6 |  |  | 50 |  |  |  |  |  |
| Saeed et al., (2023) | China | Omega-6 | MR base platform (http://www.mrbase.org) | 13506 | 8 | ADHD | Demontiset al.,2019 | 20138 | 35191 | NR |
| Davyson et al., (2023) | UK | Omega-3 | UKB | 88268 | 27 | MDD | PGC | 8840 | 20917 | 29757 |
|  |  | Omega-3% |  |  | 25 |  |  |  |  |  |
|  |  | Omega-6 |  |  | 35 |  |  |  |  |  |
|  |  | Omega-6% |  |  | 33 |  |  |  |  |  |
|  |  | Omega-6:Omega-3 |  |  | 23 |  |  |  |  |  |
| Zagkos et al., (2022) | UK | Omega-3 | Kettunen et al.,2016 | NR | 45 | AD | IGAP | 17008 | NR | 74046 |
|  |  | Omega-6 |  |  | 56 |  |  |  |  |  |
| Lord et al., (2021) | UK | Omega-3 | Kettunen et al.,2016 | 13544 | 5 | AD | ADNI | 553 | NR | 886 |
|  |  |  |  |  |  |  | ANM | 330 | NR | 648 |
|  |  |  |  |  |  |  | GERAD | 2225 | NR | 3191 |
| R Carnegieet al., (2024) | UK | Omega-3 | UKB | 115078 | 43 | MDD | PGC | 135458 | 344901 | 480359 |
|  |  | Omega-3% |  |  | 33 |  |  |  |  |  |
|  |  | Omega-6 |  |  | 50 |  |  |  |  |  |
| Tianyuan Lu al., (2024) | Canada | Omega-3 | CLSA cohort | 8299 | 38 | BD | PGC | 41917 | 371549 | NR |
|  |  | Omega-6 |  |  | 56 |  |  |  |  |  |
|  |  | Omega-3% |  |  | 29 |  |  |  |  |  |
|  |  | Omega-6% |  |  | 48 |  |  |  |  |  |
|  |  | Omega-6:Omega-3 |  |  | 30 |  |  |  |  |  |

AD, Alzheimer's disease; ADHD,attention deficit hyperactivity disorder; ASD,autism spectrum disorder; BD,bipolar disorder; IVs, instrumental variables; IVW,Inverse variance weighted; MDD,major depressive disorder; NR,Not reported; OCD,obsessive-compulsive disorder; PTSD, post-traumatic stress disorder; SCZ, schizophrenia; SNP,single-nucleotide polymorphism

Table S5 Quality assessment results.

|  | **Zhang et al., (2023)** | **Xu et al., (2023)** | **Milaneschi et al., (2019)** | **Ma et al., (2023)** | **Zeng et al., (2022)** | **Rebecca et al., (2022)** | **Saeed et al., (2023)** | **Davyson et al., (2023)** | **Zagkos et al., (2022)** | **Lord et al., (2021)** | **R Carnegieet al., (2024)** | **Tianyuan Lu al., (2024)** |
| --- | --- | --- | --- | --- | --- | --- | --- | --- | --- | --- | --- | --- |
| **Title and Abstract** |  |  |  |  |  |  |  |  |  |  |  |  |
| **Objectives** |  |  |  |  |  |  |  |  |  |  |  |  |
| **Study Design and Data Sources** |  |  |  |  |  |  |  |  |  |  |  |  |
| **MR Assumption 1** |  |  |  |  |  |  |  |  |  |  |  |  |
| **MR Assumption 2** |  |  |  |  |  |  |  |  |  |  |  |  |
| **MR Assumption 3** |  |  |  |  |  |  |  |  |  |  |  |  |
| **Analytical Methods** |  |  |  |  |  |  |  |  |  |  |  |  |
| **Sensitivity and Additional Analyses** |  |  |  |  |  |  |  |  |  |  |  |  |
| **Participant Summary Statistics** |  |  |  |  |  |  |  |  |  |  |  |  |
| **Main MR Results** |  |  |  |  |  |  |  |  |  |  |  |  |
| **Addresses Limitations** |  |  |  |  |  |  |  |  |  |  |  |  |
| **Interprets MR Results** |  |  |  |  |  |  |  |  |  |  |  |  |
| **Power Calculations** |  |  |  |  |  |  |  |  |  |  |  |  |
| **Discusses Generalizability of Results** |  |  |  |  |  |  |  |  |  |  |  |  |

For quality assessment, information was extracted based on a template developed from the Strengthening the Reporting of Observational Studies in Epidemiology using Mendelian Randomization (STROBE-MR) guidelines.In the quality assessment table, green represents that the study has fully addressed the specific aspect, with high quality and rigorous methodology, ensuring reliable results. Yellow indicates that the study has partially addressed the aspect, but lacks sufficient detail or clarity, suggesting some uncertainty or potential limitations. Red signifies that the study has either not addressed the aspect at all or has done so poorly, resulting in a significant risk of bias and lower overall reliability.

Table S6 De novo Mendelian Randomization meta-analysis and integration with existing research findings.

| Disease | **First author, year** | **Omega** | | | | | **Omega (%)** | | | | |
| --- | --- | --- | --- | --- | --- | --- | --- | --- | --- | --- | --- |
|  |  | **I²**（**%**） | **N** | **OR** | **Lower** | **Upper** | **I²**（**%**） | **N** | **OR** | **Lower** | **Upper** |
| **Omega-3** |  |  |  |  |  |  |  |  |  |  |  |
| AD | **De novo MR meta** | 0 | 3 | 1.058 | 0.986 | 1.13 | 0 | 2 | 1.04 | 0.975 | 1.105 |
|  | Zagkos et al., (2022) | NA | 1 | 1.431 | 0.640 | 3.199 | NA | 0 | NA | NA | NA |
|  | Lord et al., (2021) | NA | 1 | 1.061 | 0.936 | 1.203 | NA | 0 | NA | NA | NA |
| ADHD | **De novo MR meta** | 0 | 12 | 0.948 | 0.92 | 0.977 | 0 | 5 | 0.956 | 0.905 | 1.006 |
|  | Xu et al., (2023) | NA | 0 | NA | NA | NA | NA | 1 | 0.893 | 0.765 | 1.021 |
| Anxiety | **De novo MR meta** | 0 | 4 | 0.967 | 0.942 | 0.993 | 0 | 4 | 0.962 | 0.937 | 0.986 |
|  | Xu et al., (2023) | NA | 0 | NA | NA | NA | NA | 1 | 1.193 | 0.946 | 1.440 |
| ASD | **De novo MR meta** | 0 | 4 | 0.995 | 0.924 | 1.066 | 0 | 2 | 1.009 | 0.916 | 1.103 |
|  | Xu et al., (2023) | NA | 0 | NA | NA | NA | NA | 1 | 0.958 | 0.807 | 1.110 |
| BD | **De novo MR meta** | 0 | 2 | 0.875 | 0.822 | 0.928 | 0 | 0 | NA | NA | NA |
|  | Zhang et al., (2023) | NA | 1 | 0.884 | 0.796 | 0.982 | NA | 0 | NA | NA | NA |
|  | Tianyuan Lu al., (2024) | NA | 1 | 0.933 | 0.863 | 1.008 | NA | 1 | 0.887 | 0.830 | 0.947 |
| Depression | **De novo MR meta** | 0 | 3 | 0.998 | 0.99 | 1.005 | 0 | 2 | 0.99 | 0.983 | 0.997 |
|  | Zeng et al., (2022) | NA | 1 | 0.956 | 0.911 | 1.004 | NA | 0 | NA | NA | NA |
| Mania | **De novo MR meta** | 0 | 3 | 0.998 | 0.856 | 1.139 | 0 | 2 | 0.935 | 0.797 | 1.073 |
| MDD | **De novo MR meta** | 24.9 | 22 | 0.932 | 0.906 | 0.957 | 22.6 | 18 | 0.928 | 0.907 | 0.949 |
|  | Ma et al., (2023) Xu et al., (2023) | NA | 1 | 0. 957 | 0. 875 | 1. 047 | NA | 1 | 0.953 | 0.913 | 0.994 |
|  | Rebecca et al., (2022) | NA | 1 | 0.960 | 0.930 | 0.980 | NA | 1 | 0.960 | 0.930 | 0.980 |
|  | Davyson et al., (2023) | NA | 1 | 0.955 | 0.930 | 0.980 | NA | 1 | 0.948 | 0.919 | 0.976 |
|  | R Carnegieet al., (2024) | NA | 1 | 0.960 | 0.930 | 0.980 | NA | 1 | 0.960 | 0.930 | 0.980 |
| OCD | **De novo MR meta** | 2.4 | 3 | 0.891 | 0.798 | 0.984 | 0 | 2 | 0.905 | 0.81 | 1 |
|  | Xu et al., (2023) | NA | 0 | NA | NA | NA | NA | 1 | 1.063 | 0.809 | 1.317 |
| PsD | **De novo MR meta** | 0 | 29 | 0.94 | 0.907 | 0.974 | 0 | 19 | 0.911 | 0.878 | 0.945 |
| PTSD | **De novo MR meta** | 0 | 3 | 1.026 | 0.927 | 1.124 | 0 | 1 | 1 | 0.855 | 1.145 |
|  | Xu et al., (2023) | NA | 0 | NA | NA | NA | NA | 1 | 0.970 | 0.869 | 1.071 |
| SCZ | **De novo MR meta** | 4.3 | 13 | 0.95 | 0.904 | 0.997 | 0 | 10 | 1.029 | 0.976 | 1.083 |
|  | Xu et al., (2023) | NA | 0 | NA | NA | NA | NA | 1 | 1.014 | 0.900 | 1.127 |
| **Omega-6** |  |  |  |  |  |  |  |  |  |  |  |
| AD | **De novo MR meta** | 0 | 2 | 1.17 | 0.953 | 1.387 | 0 | 2 | 1.025 | 0.901 | 1.149 |
|  | Zagkos et al., (2022) | NA | 1 | 1.737 | 0.972 | 3.106 | NA | 0 | NA | NA | NA |
| ADHD | **De novo MR meta** | 0 | 12 | 0.928 | 0.896 | 0.96 | 0 | 6 | 0.958 | 0.884 | 1.032 |
|  | Saeed et al., (2023) Xu et al., (2023) | NA | 1 | 0.940 | 0.860 | 1.020 | NA | 1 | 0.995 | 0.944 | 1.047 |
| Anxiety | **De novo MR meta** | 0 | 4 | 1.025 | 0.989 | 1.062 | 0 | 3 | 1.033 | 0.984 | 1.083 |
|  | Xu et al., (2023) | NA | 0 | NA | NA | NA | NA | 1 | 0.966 | 0.728 | 1.204 |
| ASD | **De novo MR meta** | 0 | 5 | 1.008 | 0.949 | 1.068 | 0 | 4 | 0.958 | 0.888 | 1.027 |
|  | Xu et al., (2023) | NA | 0 | NA | NA | NA | NA | 1 | 0.971 | 0.859 | 1.083 |
| BD | **De novo MR meta** | 0 | 3 | 0.941 | 0.873 | 1.008 | 0 | 0 | NA | NA | NA |
|  | Zhang et al., (2023) | NA | 1 | 1.049 | 0.902 | 1.221 | NA | 0 | NA | NA | NA |
|  | Tianyuan Lu al., (2024) | NA | 1 | 1.060 | 0.993 | 1.131 | NA | 1 | 0.969 | 0.877 | 1.071 |
| Depression | **De novo MR meta** | 0 | 7 | 1 | 0.996 | 1.004 | 0 | 1 | 0.99 | 0.98 | 1 |
|  | Zeng et al., (2022) | NA | 1 | 1.007 | 0.944 | 1.073 | NA | 0 | NA | NA | NA |
| Mania | **De novo MR meta** | 0 | 3 | 0.947 | 0.77 | 1.124 | 0 | 2 | 0.805 | 0.602 | 1.008 |
| MDD | **De novo MR meta** | 0 | 26 | 0.993 | 0.975 | 1.012 | 0 | 12 | 1.005 | 0.977 | 1.033 |
|  | Rebecca et al., (2022) Xu et al., (2023) | NA | 1 | 1.010 | 0.970 | 1.050 | NA | 1 | 0.989 | 0.958 | 1.019 |
|  | Davyson et al., (2023) | NA | 1 | 0.987 | 0.954 | 1.021 | NA | 1 | 1.015 | 0.980 | 1.050 |
|  | R Carnegieet al., (2024) | NA | 1.000 | 1.010 | 0.970 | 1.050 | NA | 0 | NA | NA | NA |
| OCD | **De novo MR meta** | 0 | 3 | 1.022 | 0.899 | 1.145 | 0 | 2 | 1.225 | 1.013 | 1.437 |
|  | Xu et al., (2023) | NA | 0 | NA | NA | NA | NA | 1 | 1.001 | 0.714 | 1.288 |
| PsD | **De novo MR meta** | 0 | 30 | 1.016 | 0.973 | 1.059 | 0 | 18 | 0.910 | 0.848 | 0.973 |
| PTSD | **De novo MR meta** | 0 | 2 | 1.087 | 0.919 | 1.255 | 0 | 2 | 1.026 | 0.853 | 1.199 |
|  | Xu et al., (2023) | NA | 0 | NA | NA | NA | NA | 1 | 1.020 | 0.921 | 1.119 |
| SCZ | **De novo MR meta** | 0 | 21 | 1.001 | 0.972 | 1.029 | 0 | 11 | 0.995 | 0.944 | 1.047 |
|  | Xu et al., (2023) | NA | 0 | NA | NA | NA | NA | 1 | 1.015 | 0.909 | 1.121 |
| **Omega-6**：**Omega-3** |  |  |  |  |  |  |  |  |  |  |  |
| AD | **De novo MR meta** | 0 | 2 | 0.96 | 0.893 | 1.027 | - | - | - | - | - |
| ADHD | **De novo MR meta** | 0 | 5 | 1.049 | 0.999 | 1.099 | - | - | - | - | - |
|  | Xu et al., (2023) | NA | 1 | 1.120 | 1.010 | 1.229 | - | - | - | - | - |
| Anxiety | **De novo MR meta** | 0 | 3 | 1.045 | 1.015 | 1.076 | - | - | - | - | - |
|  | Xu et al., (2023) | NA | 1 | 0.884 | 0.669 | 1.100 | - | - | - | - | - |
| ASD | **De novo MR meta** | 0 | 3 | 0.988 | 0.919 | 1.057 | - | - | - | - | - |
|  | Xu et al., (2023) | NA | 1 | 1.032 | 0.894 | 1.169 | - | - | - | - | - |
| BD | **De novo MR meta** | 0 | 1 | 1.14 | 1.03 | 1.25 | - | - | - | - | - |
|  | Zhang et al., (2023) | NA | 1 | 1.172 | 1.046 | 1.314 | - | - | - | - | - |
|  | Xu et al., (2023) | NA | 1 | 1.042 | 0.940 | 1.143 | - | - | - | - | - |
|  | Tianyuan Lu al., (2024) | NA | 1 | 1.106 | 1.021 | 1.198 | - | - | - | - | - |
| Depression | **De novo MR meta** | 0 | 4 | 1.01 | 1.005 | 1.015 | - | - | - | - | - |
| Mania | **De novo MR meta** | 0 | 2 | 1.01 | 0.856 | 1.164 | - | - | - | - | - |
| MDD | **De novo MR meta** | 44 | 17 | 1.086 | 1.052 | 1.12 | - | - | - | - | - |
|  | Xu et al., (2023) | NA | 1 | 1.029 | 0.992 | 1.066 | - | - | - | - | - |
|  | Davyson et al., (2023) | NA | 1 | 1.054 | 1.023 | 1.085 | - | - | - | - | - |
| OCD | **De novo MR meta** | 0 | 2 | 1.11 | 0.978 | 1.242 | - | - | - | - | - |
|  | Xu et al., (2023) | NA | 1 | 0.982 | 0.736 | 1.228 | - | - | - | - | - |
| PsD | **De novo MR meta** | 0 | 20 | 1.075 | 1.033 | 1.117 | - | - | - | - | - |
| PTSD | **De novo MR meta** | 0 | 0 | NA | NA | NA | - | - | - | - | - |
| SCZ | **De novo MR meta** | 0 | 10 | 1.007 | 0.965 | 1.048 | - | - | - | - | - |
|  | Xu et al., (2023) | NA | 1 | 0.959 | 0.824 | 1.095 | - | - | - | - | - |

| **Disease** | **First author, year** | **Omega** | | | | | **Omega (%)** | | | | |
| --- | --- | --- | --- | --- | --- | --- | --- | --- | --- | --- | --- |
|  |  | **I²**（**%**） | **N** | **OR** | **Lower** | **Upper** | **I²**（**%**） | **N** | **OR** | **Lower** | **Upper** |
| **Omega-3** |  |  |  |  |  |  |  |  |  |  |  |
| PsD-anankastic | **De novo MR meta** | 0 | 3 | 1.015 | 0.872 | 1.159 | 0 | 2 | 0.93 | 0.801 | 1.059 |
| PsD-Anxietyious | **De novo MR meta** | 0 | 3 | 1.071 | 0.876 | 1.266 | 0 | 2 | 0.964 | 0.781 | 1.148 |
| PsD-dependent | **De novo MR meta** | 0 | 3 | 0.777 | 0.646 | 0.909 | 0 | 2 | 0.805 | 0.66 | 0.95 |
| PsD-dissocial | **De novo MR meta** | 0 | 3 | 1.019 | 0.819 | 1.22 | 0 | 2 | 0.994 | 0.787 | 1.2 |
| PsD-emotional | **De novo MR meta** | 0 | 3 | 0.911 | 0.849 | 0.973 | 0 | 2 | 0.88 | 0.818 | 0.942 |
| PsD-histrionic | **De novo MR meta** | 0 | 3 | 1.130 | 0.651 | 1.609 | 0 | 1 | 1.000 | 0.46 | 1.54 |
| PsD-other | **De novo MR meta** | 0 | 3 | 0.939 | 0.863 | 1.015 | 0 | 2 | 0.915 | 0.837 | 0.993 |
| PsD-paranoid | **De novo MR meta** | 0 | 3 | 0.974 | 0.791 | 1.157 | 0 | 2 | 1.016 | 0.815 | 1.218 |
| PsD-schizoid | **De novo MR meta** | 0 | 3 | 0.994 | 0.814 | 1.175 | 0 | 2 | 1.088 | 0.882 | 1.295 |
| **Omega-6** |  |  |  |  |  |  |  |  |  |  |  |
| PsD-anankastic | **De novo MR meta** | 0 | 3 | 1.088 | 0.889 | 1.288 | 0 | 1 | 0.98 | 0.58 | 1.38 |
| PsD-Anxietyious | **De novo MR meta** | 0 | 3 | 1.103 | 0.852 | 1.355 | 0 | 2 | 0.773 | 0.519 | 1.027 |
| PsD-dependent | **De novo MR meta** | 0 | 3 | 0.807 | 0.626 | 0.989 | 0 | 2 | 1.099 | 0.732 | 1.466 |
| PsD-dissocial | **De novo MR meta** | 0 | 3 | 0.962 | 0.721 | 1.203 | 0 | 2 | 0.838 | 0.538 | 1.138 |
| PsD-emotional | **De novo MR meta** | 0 | 3 | 1.035 | 0.948 | 1.123 | 0 | 1 | 0.93 | 0.745 | 1.115 |
| PsD-histrionic | **De novo MR meta** | 0 | 3 | 1.372 | 0.725 | 2.020 | 0 | 2 | 1.103 | 0.337 | 1.869 |
| PsD-other | **De novo MR meta** | 0 | 2 | 1.018 | 0.913 | 1.123 | 0 | 2 | 1.065 | 0.906 | 1.224 |
| PsD-paranoid | **De novo MR meta** | 37.4 | 2 | 0.974 | 0.658 | 1.29 | 0 | 2 | 1.005 | 0.661 | 1.35 |
| PsD-schizoid | **De novo MR meta** | 50.1 | 3 | 0.847 | 0.567 | 1.127 | 0 | 2 | 1.639 | 1.04 | 2.238 |
| **Omega-6**：**Omega-3** |  |  |  |  |  |  |  |  |  |  |  |
| PsD-anankastic | **De novo MR meta** | 0 | 2 | 1.06 | 0.903 | 1.217 | - | - | - | - | - |
| PsD-Anxietyious | **De novo MR meta** | 0 | 2 | 1 | 0.806 | 1.194 | - | - | - | - | - |
| PsD-dependent | **De novo MR meta** | 0 | 2 | 1.29 | 1.053 | 1.527 | - | - | - | - | - |
| PsD-dissocial | **De novo MR meta** | 0 | 2 | 1.022 | 0.802 | 1.243 | - | - | - | - | - |
| PsD-emotional | **De novo MR meta** | 0 | 2 | 1.12 | 1.037 | 1.203 | - | - | - | - | - |
| PsD-histrionic | **De novo MR meta** | 0 | 2 | 0.843 | 0.496 | 1.189 | - | - | - | - | - |
| PsD-other | **De novo MR meta** | 0 | 3 | 1.065 | 0.970 | 1.16 | - | - | - | - | - |
| PsD-paranoid | **De novo MR meta** | 0 | 3 | 0.975 | 0.759 | 1.192 | - | - | - | - | - |
| PsD-schizoid | **De novo MR meta** | 0 | 2 | 0.964 | 0.747 | 1.181 | - | - | - | - | - |

Red numbers indicate positive effects. Blue numbers indicate negative effects. AD, Alzheimer's disease; ADHD, attention deficit hyperactivity disorder; ASD, autism spectrum disorder; BD, bipolar disorder; MDD, major depressive disorder; MR, Mendelian randomization; OR, odds ratio; OCD, obsessive-compulsive disorder; PTSD, post-traumatic stress disorder; PsD, personality disorder; PsD-anankastic, anankastic personality disorder; PsD-anxious, anxious personality disorder; PsD-dependent, dependent personality disorder; PsD-dissocial, dissocial personality disorder; PsD-emotional, emotionally unstable personality disorder; PsD-histrionic, histrionic personality disorder; PsD-other, mixed and other personality disorders; PsD-paranoid, paranoid personality disorder; PsD-schizoid, schizoid personality disorder; PUFAs, polyunsaturated fatty acids; SCZ, schizophrenia.

Table S7 Positive causal relationship between PUFAs and mental disorders through De Novo Mendelian randomization.

| **Exposure** | **Exposure ID** | **Outcome** | **Outcome ID** | **N snp** | **P** | **Effect size with 95%CI** |
| --- | --- | --- | --- | --- | --- | --- |
| Omega-3 | met-c-855 | OCD | F5_0CD | 6 | 0.046 | 0.764 [0.560,0.969] |
|  | ebi-a-GCST90092931 | MDD | ebi-a-GCST90086061 | 41 | 0.006 | 0.789 [0.656,0.922] |
|  | met-d-0mega_3 | MDD | ebi-a-GCST90086061 | 40 | 0.011 | 0.811 [0.678,0.943] |
|  | ebi-a-GCST90092931 | MDD | ebi-a-GCST90086059 | 41 | 0.007 | 0.874 [0.788,0.960] |
|  | met-d-0mega_3 | BD | F5_BIPO | 46 | 0.002 | 0.874 [0.799,0.950] |
|  | ebi-a-GCST90092931 | BD | F5_BIPO | 46 | 0.006 | 0.883 [0.805,0.961] |
|  | ebi-a-GCST90092931 | MDD | ebi-a-GCST90086062 | 41 | 0.004 | 0.885 [0.810,0.959] |
|  | met-d-0mega_3 | MDD | ebi-a-GCST90086059 | 40 | 0.018 | 0.888 [0.800.0.975] |
|  | ebi-a-GCST90092931 | MDD | ebi-a-GCST90086058 | 41 | 0.042 | 0.890 [0.789,0.990] |
|  | met-d-0mega_3 | MDD | ebi-a-GCST90086062 | 40 | 0.014 | 0.900 [0.825,0.976] |
|  | met-c-855 | SCZ | ieu-b-5099 | 6 | 0.039 | 0.919 [0.845,0.993] |
| Omega-3% | met-d-Omega_3_pct | MDD | ebi-a-GCST90086058 | 29 | 0.014 | 0.869 [0.772,0.967] |
|  | ebi-a-GCST90092932 | MDD | ebi-a-GCST90086059 | 27 | 0.009 | 0.879 [0.795,0.964] |
|  | met-d-Omega_3_pct | MDD | ebi-a-GCST90086059 | 29 | 0.014 | 0.881 [0.791,0.970] |
|  | met-d-Omega_3_pct | PsD-emotional | F5_EMOPER | 35 | 0.012 | 0.882 [0.795,0.969] |
|  | ebi-a-GCST90092932 | PsD-emotional | F5_EMOPER | 31 | 0.014 | 0.885 [0.798,0.972] |
|  | met-d-Omega_3_pct | MDD | ebi-a-GCST90086062 | 29 | 0.004 | 0.885 [0.811,0.960] |
|  | met-d-Omega_3_pct | MDD | ebi-a-GCST009979 | 38 | 0.003 | 0.922 [0.873,0.971] |
|  | met-d-Omega_3_pct | Anxiety | F5_ALLAnxietyIOUS | 35 | 0.050 | 0.959 [0.918,0.999] |
|  | ebi-a-GCST90092932 | Depression | 20544-11 | 35 | 0.002 | 0.988 [0.981,0.996] |
|  | ebi-a-GCST90092932 | Depression | 20544-11 | 35 | 0.002 | 1.012 [1.004,1.019] |
|  | ebi-a-GCST90092932 | MDD | ebi-a-GCST009979 | 33 | 0.034 | 1.068 [1.003,1.134] |
|  | ebi-a-GCST90092932 | MDD | ebi-a-GCST90086062 | 27 | 0.010 | 1.115 [1.022,1.208] |
|  | ebi-a-GCST90092932 | MDD | ebi-a-GCST90086061 | 27 | 0.018 | 1.231 [1.019,1.443] |
| Omega-6 | met-d-0mega_6 | ADHD | eu-a-1183 | 46 | 0.011 | 0.896 [0.820,0.972] |
|  | ebi-a-GCST90092933 | ADHD | ieu-a-1183 | 47 | 0.031 | 0.910 [0.832,0.988] |
| Omega-6% | ebi-a-GCST90092935 | PsD | finn-b-KRA_PSY_PERSON | 49 | 0.014 | 0.834 [0.713,0.955] |
|  | met-d-0mega_6_pct | PsD | finn-b-KRA_PSY_PERSON | 48 | 0.038 | 0.858 [0.733,0.982] |
| Omega-6:Omega-3 | ebi-a-GCST90092934 | Depression | 20544-11 | 32 | 0.033 | 1.010 [1.001,1.019] |
|  | ebi-a-GCST90092934 | Depression | 20544-11 | 32 | 0.031 | 1.010 [1.001,1.019] |
|  | met-d-Omega 6 by_Omega_3 | Depression | 20544-11 | 38 | 0.023 | 1.010 [1.001,1.019] |
|  | met-d-Omega 6 by_Omega_3 | Depression | 20544-11 | 38 | 0.021 | 1.010 [1.001,1.019] |
|  | met-d-Omega 6 by_Omega_3 | PsD-emotional | F5_EMOPER | 32 | 0.023 | 1.132 [1.011,1.252] |
|  | ebi-a-GCST90092934 | MDD | ebi-a-GCST90086062 | 25 | 0.011 | 1.134 [1.024,1.243] |
|  | ebi-a-GCST90092934 | MDD | ebi-a-GCST90086058 | 25 | 0.037 | 1.137 [1.000,1.275] |
|  | met-d-Omega 6 by_Omega_3 | BD | F5_BIPO | 32 | 0.008 | 1.138 [1.029,1.247] |
|  | ebi-a-GCST90092934 | MDD | ebi-a-GCST90086059 | 25 | 0.004 | 1.153 [1.040,1.265] |
|  | met-d-Omega 6 by_Omega_3 | MDD | ebi-a-GCST90086059 | 32 | 0.003 | 1.157 [1.044,1.271] |
|  | met-d-Omega 6 by_Omega_3 | MDD | ebi-a-GCST90086062 | 32 | 0.003 | 1.159 [1.047,1.272] |
|  | met-d-Omega 6 by_Omega_3 | MDD | ebi-a-GCST90086058 | 32 | 0.007 | 1.191 [1.039,1.344] |
|  | met-d-Omega 6 by_Omega_3 | MDD | ebi-a-GCST90086061 | 32 | 0.018 | 1.245 [1.018,1.472] |
|  | ebi-a-GCST90092934 | MDD | ebi-a-GCST90086061 | 25 | 0.008 | 1.266 [1.043,1.489] |
|  | met-d-Omega 6 by_Omega_3 | Anxiety | finn-b-KRA_PSY_AnxietyIETY | 36 | 0.046 | 1.743 [1.648,1.838] |

ADHD, attention deficit hyperactivity disorder; BD, bipolar disorder; MDD, major depressive disorder; OCD, obsessive-compulsive disorder; PsD, personality disorder; PsD-emotional, emotionally unstable personality disorder; PUFAs, polyunsaturated fatty acids; SCZ, schizophrenia

Table S8 Search strategy

**2024.10.01**

**1. PubMed**

#1 (((((((((((((((((((((((((((((((((((((((((((((((((((((((((((Fatty Acids, Unsaturated[Title/Abstract]) OR (Acids, Unsaturated Fatty[Title/Abstract])) OR (Unsaturated Fatty Acids[Title/Abstract])) OR (Unsaturated Fatty Acid[Title/Abstract])) OR (Acid, Unsaturated Fatty[Title/Abstract])) OR (Fatty Acid, Unsaturated[Title/Abstract])) OR (Polyunsaturated Fatty Acids[Title/Abstract])) OR (Acids, Polyunsaturated Fatty[Title/Abstract])) OR (Fatty Acids, Polyunsaturated[Title/Abstract])) OR (Polyunsaturated Fatty Acid[Title/Abstract])) OR (Acid, Polyunsaturated Fatty[Title/Abstract])) OR (Fatty Acid, Polyunsaturated[Title/Abstract])) OR (Fatty Acids, Omega-3[Title/Abstract])) OR (Omega-3 Fatty Acid[Title/Abstract])) OR (Acid, Omega-3 Fatty[Title/Abstract])) OR (Fatty Acid, Omega-3[Title/Abstract])) OR (Omega 3 Fatty Acid[Title/Abstract])) OR (Omega-3 Fatty Acids[Title/Abstract])) OR (n-3 Oil[Title/Abstract])) OR (Oil, n-3[Title/Abstract])) OR (n 3 Oil[Title/Abstract])) OR (n3 Oil[Title/Abstract])) OR (Oil, n3[Title/Abstract])) OR (n-3 Fatty Acids[Title/Abstract])) OR (n 3 Fatty Acids[Title/Abstract])) OR (Omega 3 Fatty Acids[Title/Abstract])) OR (n-3 PUFA[Title/Abstract])) OR (PUFA, n-3[Title/Abstract])) OR (n 3 PUFA[Title/Abstract])) OR (n3 Fatty Acid[Title/Abstract])) OR (Fatty Acid, n3[Title/Abstract])) OR (n3 PUFA[Title/Abstract])) OR (PUFA, n3[Title/Abstract])) OR (n3 Polyunsaturated Fatty Acid[Title/Abstract])) OR (n3 Oils[Title/Abstract])) OR (n-3 Oils[Title/Abstract])) OR (n 3 Oils[Title/Abstract])) OR (N-3 Fatty Acid[Title/Abstract])) OR (Acid, N-3 Fatty[Title/Abstract])) OR (Fatty Acid, N-3[Title/Abstract])) OR (N 3 Fatty Acid[Title/Abstract])) OR (n-3 Polyunsaturated Fatty Acid[Title/Abstract])) OR (n 3 Polyunsaturated Fatty Acid[Title/Abstract])) OR (Fatty Acids, Omega-6[Title/Abstract])) OR (Acids, Omega-6 Fatty[Title/Abstract])) OR (Omega-6 Fatty Acid[Title/Abstract])) OR (Acid, Omega-6 Fatty[Title/Abstract])) OR (Fatty Acid, Omega-6[Title/Abstract])) OR (Omega 6 Fatty Acid[Title/Abstract])) OR (Omega-6 Fatty Acids[Title/Abstract])) OR (Omega 6 Fatty Acids[Title/Abstract])) OR (N-6 Fatty Acid[Title/Abstract])) OR (Acid, N-6 Fatty[Title/Abstract])) OR (Fatty Acid, N-6[Title/Abstract])) OR (N 6 Fatty Acid[Title/Abstract])) OR (Fatty Acids, Omega 6[Title/Abstract])) OR (N-6 Fatty Acids[Title/Abstract])) OR (Acids, N-6 Fatty[Title/Abstract])) OR (Fatty Acids, N-6[Title/Abstract])) OR (N 6 Fatty Acids[Title/Abstract]))

#2 (Mendelian Randomization[Title/Abstract]) OR (Analysis, Mendelian Randomization[Title/Abstract]))

#3 #1 AND #2

154 articles were retrieved

**2. Embase**

#1 'unsaturated fatty acid'/exp OR 'alkenyl fatty acid' OR 'fats, unsaturated' OR 'fatty acid, unsaturated' OR 'fatty acids, unsaturated' OR 'UFA' OR 'unsaturated fat' OR 'unsaturated lipid' OR 'unsaturated fatty acid'

#2 'omega 3 fatty acid'/exp OR 'bilantin omega' OR 'conchol 36' OR 'eicosa e' OR 'eicosapen' OR 'epaisdin' OR 'epanova' OR 'fatty acids, omega 3' OR 'fatty acids, omega-3' OR 'n 3 fatty acid' OR 'n 3 polyunsaturated fatty acid' OR 'omega 3' OR 'omega 3 carboxylic acid' OR 'omega 3 carboxylic acids' OR 'omega 3 feingold' OR 'omega 3 plus' OR 'omega 3 polyunsaturated fatty acid' OR 'omega forte' OR 'omega-3-carboxylic acids' OR 'omega3 polyunsaturated fatty acid' OR 'sakana' OR 'sanhelios omega 3' OR 'omega 3 fatty acid'

#3'omega 6 fatty acid'/exp OR 'fatty acids, omega 6' OR 'fatty acids, omega-6' OR 'n 6 fatty acid' OR 'omega 6 polyunsaturated fatty acid' OR 'omega 6 fatty acid'

#4'Mendelian randomization analysis'/exp OR 'Mendelian randomisation' OR 'Mendelian randomization' OR 'Mendelian randomization analysis'

#5 #1 OR #2 OR #3 AND #4

382 articles were retrieved

**3. Cochrane Library**

#1 MeSH descriptor: [Fatty Acids, Unsaturated] explode all trees

#2 (Acids, Unsaturated Fatty OR Unsaturated Fatty Acids OR Unsaturated Fatty Acid OR Acid, Unsaturated Fatty OR Fatty Acid, Unsaturated OR Polyunsaturated Fatty Acids OR Acids, Polyunsaturated Fatty OR Fatty Acids, Polyunsaturated OR Polyunsaturated Fatty Acid OR Acid, Polyunsaturated Fatty OR Fatty Acid, Polyunsaturated):ti,ab,kw

#3 MeSH descriptor: [Fatty Acids, Omega-3] explode all trees

#4 (Omega-3 Fatty Acid OR Acid, Omega-3 Fatty OR Fatty Acid, Omega-3 OR Omega 3 Fatty Acid OR Omega-3 Fatty Acids OR n-3 Oil OR Oil, n-3 OR n 3 Oil OR n3 Oil OR Oil, n3 OR n-3 Fatty Acids OR n 3 Fatty Acids OR Omega 3 Fatty Acids OR n-3 PUFA OR PUFA, n-3 OR n 3 PUFA OR n3 Fatty Acid OR Fatty Acid, n3 OR n3 PUFA OR PUFA, n3 OR n3 Polyunsaturated Fatty Acid OR n3 Oils OR n-3 Oils OR n 3 Oils OR N-3 Fatty Acid OR Acid, N-3 Fatty OR Fatty Acid, N-3 OR N 3 Fatty Acid OR n-3 Polyunsaturated Fatty Acid OR n 3 Polyunsaturated Fatty Acid):ti,ab,kw

#5MeSH descriptor: [Fatty Acids, Omega-6] explode all trees

#6(Acids, Omega-6 Fatty OR Omega-6 Fatty Acid OR Acid, Omega-6 Fatty OR Fatty Acid, Omega-6 OR Omega 6 Fatty Acid OR Omega-6 Fatty Acids OR Omega 6 Fatty Acids OR N-6 Fatty Acid OR Acid, N-6 Fatty OR Fatty Acid, N-6 OR N 6 Fatty Acid OR Fatty Acids, Omega 6 OR N-6 Fatty Acids OR Acids, N-6 Fatty OR Fatty Acids, N-6 OR N 6 Fatty Acids):ti,ab,kw

#7 MeSH descriptor: [Mendelian Randomization Analysis] explode all trees

#8 (Analysis, Mendelian Randomization):ti,ab,kw

#9 #1 OR #2 OR #3 OR #4 OR #5 OR #6

#10 #7 OR #8

#11 #9 AND #10

39 articles were retrieved

575 articles in total
